# Germline *NF2* variant position constrains somatic second hits and determines clinical severity in Neurofibromatosis Type 2-related schwannomatosis

**DOI:** 10.64898/2026.07.27.26359036

**Authors:** Niveditha Ravindra, David Asuzu, Emma Celano, Santosh Kumar, Arshi Chopra, Debjani Mandal, Dustin Mullaney, Dhruval Bhatt, Maxwell T Laws, Christina Hayes, Isac Kunnath, Bayu Sisay, Abdel Elkahloun, Ashok Ashthagiri, Tanya Lehky, Christopher Zalewski, John D Heiss, Hung J Kim, Prashant Chittiboina

## Abstract

**Purpose:** Neurofibromatosis type 2-related schwannomatosis (NF2-SWN) is an autosomal dominant tumor syndrome with complete penetrance and variable expressivity. Underlying patterns of disease burden and severity are largely unexplained.

**Methods:** We comprehensively phenotyped 168 NF2-SWN patients over a mean duration of 4.5 years. We used a custom sequencing panel of *NF2* and schwannomatosis genes to identify germline (n=166) and somatic variants in tumors (n=37). An optimized composite severity (CSS) score based on clinical and radiological data was created to analyze the effect of genetic variants on phenotype.

**Results:** We found significant variable expressivity not explainable by demographic variables. Germline variants included premature termination (42%), splice-site (18%), and large deletions (16%). The CSS successfully predicted worsening clinical function in patients. Unsupervised clustering of clinical data revealed distinct phenotypic clusters that corresponded to CSS. Mosaicism, however, was not associated with CSS or any other disease severity marker. CSS was significantly associated with germline variant location along the *NF2* locus. Specifically, FERM-F1 and the α-helical variants were associated with increased disease severity. Within tumors, germline variants with severe effects on merlin acquired milder somatic second-hits at the *NF2* locus.

**Conclusion:** We identified a second-hit modifier to the Mendelian first-hit: severe germline variants were associated with milder somatic variants, and vice versa. This phenomenon partly explains the variable expressivity in NF2-SWN.

**Context Summary:** Neurofibromatosis type 2-related schwannomatosis (NF2-SWN) displays striking variability in tumor burden and clinical severity, but the biological basis for this heterogeneity has remained unclear. Using deep longitudinal phenotyping, whole-neuroaxis imaging, and integrated germline-somatic genomic analysis, we show that germline *NF2* variant position determines the allowable somatic ‘second hits’ that can give rise to tumors. These data support a merlin-dosage model in which tumorigenesis likely occurs only within an intermediate range of merlin function, bounded by insufficiently tumorigenic and lethal levels on each side. These findings shed light on long-standing inconsistencies in genotype-phenotype relationships in NF2-SWN and help provide a mechanistic basis for predicting disease severity.

## Introduction

Neurofibromatosis type 2-related schwannomatosis (NF2-SWN) is an autosomal dominant tumor-predisposition syndrome with an incidence of approximately 1 in 28,000 and prevalence of 1 in 50,000(1). NF2-SWN is characterized pathognomonically by bilateral vestibular schwannomas. Patients often have severe clinical consequences from multiple other tumors including cranial, spinal and peripheral nerve schwannomas, cranial and spinal meningiomas, spinal ependymomas, and non-neoplastic manifestations such as cataracts, peripheral neuropathy, and speech/swallowing dysfunction(2–4). Despite near-complete penetrance, NF2-SWN displays striking variable expressivity, with substantial heterogeneity in tumor burden, anatomic distribution, functional impairment, and clinical course even within families.

NF2-SWN arises from pathogenic variants in the *NF2* gene on chromosome 22q12, which encodes the tumor suppressor merlin. Merlin is a member of the ezrin-radixin-moesin (ERM) family of cytoskeletal adapter proteins and plays a central role in contact inhibition, membrane receptor signaling, and growth control. Structurally, merlin consists of an N-terminal FERM domain, a central α-helical linker, and a C-terminal tail, enabling it to act as a scaffold that links membrane proteins to intracellular signaling pathways, including receptor tyrosine kinases, PI3K/AKT, cell adhesion complexes, and the Hippo pathway(5). Loss of merlin function disrupts these processes, promoting abnormal proliferation and survival.

Tumor development in NF2-SWN conforms to the classical Knudson two-hit model, in which a germline pathogenic variant is followed by somatic inactivation of the remaining allele (6,7). We have found that tumors exhibit variable merlin dosage and activity measurable as a distinct transcriptomic signature – the merlin depletion score (8). Moreover, peripheral neuropathy is thought to arise from haploinsufficiency(9), reflecting reduced merlin dosage rather than complete loss of function. NF2-SWN patients therefore experience a combination of one-hit effects (such as neuropathy) and two-hit effects (tumors). This implies that disease severity reflects the combination of germline and somatic events, which jointly determine merlin dosage across different cell types.

Multiple studies have attempted to correlate NF2-SWN genotype with clinical severity(10–12). Prior work suggested that germline truncating variants conferred worse prognosis than missense or splice-site variants. These observations informed the development of the Genetic Severity Score (GSS)(13), which integrates germline variant type, location, and mosaic status to predict disease severity. While the GSS demonstrated predictive value for functional decline, subsequent studies failed to reproduce consistent genotype-phenotype correlations (11,14), and the GSS has limited utility for predicting tumor burden, progression, or overall clinical trajectory. These inconsistencies highlight a fundamental challenge: traditional categorical descriptors of genotype do not capture the biological complexity underlying phenotypic variability in NF2-SWN.

A limitation of prior studies is the reliance on coarse or one-dimensional measures of disease severity(10,11,13,15–17). NF2-SWN severity has been assessed using age at symptom onset, hearing loss, tumor counts, surgical history, or mortality, but no single metric adequately reflects the multidimensional nature of the disease (4). We hypothesized that deep phenotypic characterization combined with quantitative severity modeling could reveal the first principles governing NF2-SWN disease phenotype. Here, we performed comprehensive longitudinal phenotyping of a natural history cohort, integrating volumetric magnetic resonance imaging (MRI)-derived tumor burden, functional outcomes, surgical history, and germline and somatic genomic data. We developed a Composite Severity Score (CSS) that captures multidimensional disease burden and outperforms traditional one-dimensional severity metrics. Applying the CSS, we found that germline genomic position rather than variant type emerged as a primary determinant of disease severity. Further, we demonstrate that germline variant position constrains the allowable landscape of somatic second hits and only specific combinations of germline and somatic alterations give rise to viable tumors in NF2-SWN patients. Together with neuropathy analyses that reflect haploinsufficiency in non-replicating neurons, these findings support a merlin dosage model in which disease manifestations arise within narrow functional windows defined by normal function, tumor permissiveness, and cellular non-viability. This merlin dosage-based framework provides a unifying explanation for NF2-SWN phenotypic heterogeneity and reconciles long-standing inconsistencies in genotype-phenotype relationships.

## Methods

### Study design and data collection

This is a prospective, longitudinal study of patients meeting the clinical criteria for NF2-SWN with/without genetic testing. The study was approved by the Combined Neuroscience Institutional Review Board of the National Institutes of Health (IRB approval: 08-N-0044, NCT00598351). Written and informed consent was obtained from every participant prior to enrollment. MR imaging with gadolinium, audiometry and vestibular evaluation were performed at each visit. Clinicians assessed Karnofsky performance status (KPS), ambulatory status, and abbreviated ASIA motor scores. Patients were followed for up to 5 years. Additional details are provided in Sup. Methods 1.1-1.3.

### Genetic testing

CLIA-certified testing was available in 69 patients, and 125 underwent in-house targeted deep sequencing of *NF2*, *SMARCE1*, *LZTR1*, *SMARCB1*, and *SUFU* genes. Surgically removed tumors (n=37; 30 patients) were sequenced using the same panel. Germline data were analyzed with Qiagen CLC Genomics Workbench while tumor data was processed on NIH’s Biowulf computing cluster. Variants were classified per American College of Medical Genetics/Association of Molecular Pathologists guidelines and an allele frequency cutoff of <30% was used for mosaic variants. Further details are provided in the Sup. Methods 1.4.

### Construction and validation of Composite Severity Score

A retrospective baseline severity metric was constructed using three clinically relevant, minimally collinear variables-total tumor count, total tumor volume, and KPS derived from whole-neuroaxis MRI annotations and baseline clinical assessment. A forward-looking Progression Index was generated from prospective outcomes (new tumors, tumor surgeries, and change in KPS), each standardized and averaged to link baseline features with subsequent deterioration without incorporating post-enrollment data into score construction. Multiple strategies were evaluated, including equal-weight composites, Principal Component Analysis (PCA), and supervised Elastic Net and LASSO models (scikit-learn). The final CSS was derived using Elastic Net regression (ElasticNetCV) with 5-fold cross-validation. Model performance was internally validated by bootstrap resampling (refer to Sup. Methods 1.5 for full details).

### Phenotypic clustering

Tumor phenotype clustering used five craniospinal burden metrics (cranial meningiomas, cranial schwannomas, spinal meningiomas, spinal schwannomas, spinal intramedullary tumors), excluding patients with missing data. Tumor counts were z-scaled and grouped by k-means clustering, with the optimal number of clusters (k = 1-10) chosen using the elbow method. Cluster assignments were visualized by PCA (further details are provided in Sup. Methods 1.6).

### Allele impact

To identify germline-somatic interactions, we assembled a cohort of 77 tumors by combining our institutional cohort (n = 37) complemented with previous reports with complete germline and somatic mutation data.We developed a 0–1 allele impact score for *NF2* variants based on variant type, exon position, or deletion extent. K-means clustering (k=3) on paired germline-somatic scores was evaluated via silhouette coefficient, with significance assessed by a 1,000-iteration permutation test shuffling germline-somatic pairings. Fisher’s exact test assessed tumor-type enrichment across clusters (Refer to Sup. Methods 1.7-1.8).

### Statistical analysis

Associations between cluster identity and genotype categories (variant type, truncating vs non-truncating status, mosaicism, GSS, and predicted merlin domain) were tested with χ² tests. Differences in tumor-type burden across merlin domains were assessed using ANOVA followed by Tukey post-hoc tests. Relationships between phenotype clusters and the CSS were examined with violin plots and χ² tests.

## Results

### Deep longitudinal phenotyping reveals the multidimensional structure of NF2 disease

To define clinical and biological determinants of NF2-SWN severity, we conducted a comprehensive natural history study (NCT00598351) with deep longitudinal phenotyping, and multimodal imaging in 168 prospectively enrolled patients. At baseline and annual visits, participants underwent detailed clinical evaluations, volumetric brain and spine MRI, audio-vestibular testing, and biospecimen collection. Deep next-generation sequencing (NGS) was performed using a targeted panel including *NF2*, *LZTR1*, *SMARCB1*, *SMARCE1*, and *SUFU* genes(**Fig. 1A**). This panel enabled detailed assessment of pathogenic variants, including type, locus, allele fraction, and domain-level effects. We then integrated tumor metrics, clinical severity measures, genotype, and demographics into a unified analytical framework (**Fig. 1B**).

**Fig. 1.**
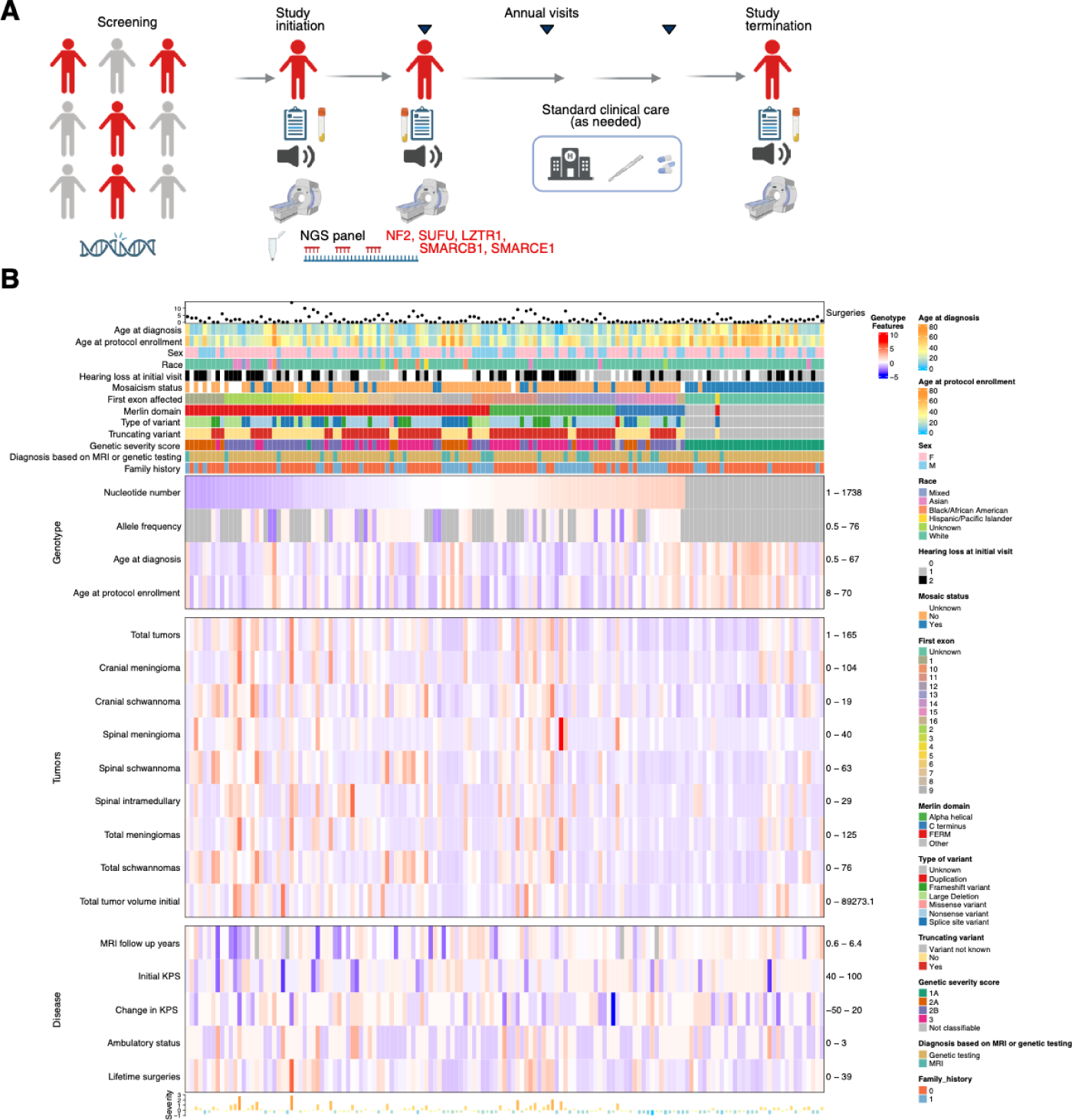
Deep phenotypic analysis reveals disease patterns in patients with NF2-SWN: **A.** A schematic summary of the natural history study. Study initiation and subsequent visits comprise clinical exams, serum collection, polymorphonuclear cells (PBMC) collection, audio-vestibular testing, and volumetric magnetic resonance imaging (MRI) of the brain and spine. Germline genotype testing was performed on PBMCs using a custom next-generation testing (NGS) for *NF2*, *LZTR1*, *SMARCB1*, *SMARCE1*, and *SUFU* genes. **B.** A summary heatmap of the natural history study data. Each column represents a patient. Patients are ordered from left to right based on the germline first affected nucleotide position in the *NF2* gene. The top annotation rows represent categorical demographic (age and sex), categorical genotypic *NF2* data (merlin domain, truncating variant, type of variant, and the predicted genetic severity score), and the relevant phenotypic information. Scaled quantitative data is represented in the main body of the heatmap. The ranges for each of the rows are given to the right of the respective row. The top sub-group of rows maps quantitative information with predictive potency. The middle sub-group of rows maps the prevalence of each of the tumor type, and total tumor volumes. The bottom sub-group of rows represents the phenotypic clinical data. The Composite severity score for each patient is plotted as a bar plot in the bottom row.

We summarized quantitative tumor metrics including cranial and spinal meningiomas and schwannomas, intramedullary tumors, and total tumor volume. We found extensive variability in tumor number, size, and distribution across patients, even within shared genotypes. Clinical severity measures included lifetime surgeries, ambulatory status, baseline KPS, and change in KPS over time. This high-dimensional dataset provided the basis for subsequent analyses: characterizing phenotypic variability (**Fig. 2**), evaluating existing genetic prediction scores (15), deriving a composite severity score (**Fig. 3**), clustering NF2-SWN phenotypes (**Fig. 4**), and investigating mechanistic links between germline variant position, somatic second hits, and disease severity (**Figs. 5–7**)(16).

**Fig. 2:**
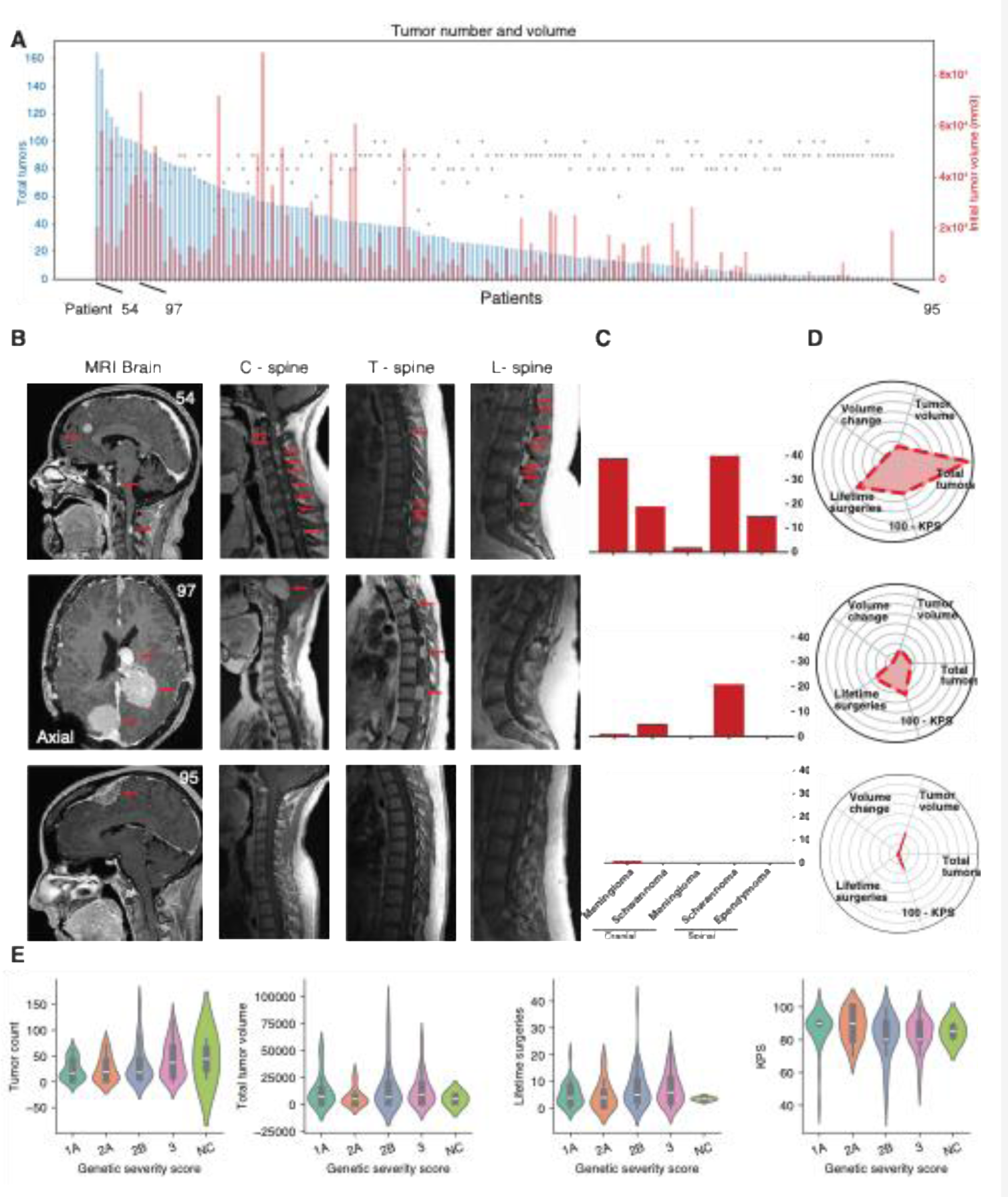
Multidimensional clinical and imaging features reveal broad phenotypic variability in NF2-SWN patients. **A.** A plot of total tumor count (blue bars), and the total tumor volume (red bars) at study initiation. Karnofsky performance status (black points) are overlaid on the tumor phenotype. **B.** MRI images (T1-weighted, post-gadolinium contrast) of representative patients (Pt54, Pt97, and Pt95). Representative slices of MRI brain, cervical spine, thoracic spine, and lumbar spine are presented, with red arrows marking detectable tumors. **C.** Bar plots summarizing the number of tumors found in each of the patients in B. **D.** Radar plots illustrating the multidimensional NF2-SWN phenotype for each representative patient. The radar plots incorporate total tumor count, total tumor volume, percent volume change during the study, number of lifetime surgeries, and functional status (100 – KPS). A larger area qualitatively reflects more severe disease. **E.** Violin plots demonstrating the relationship of individual markers of disease severity with the predictive genetic severity score.

**Fig. 3:**
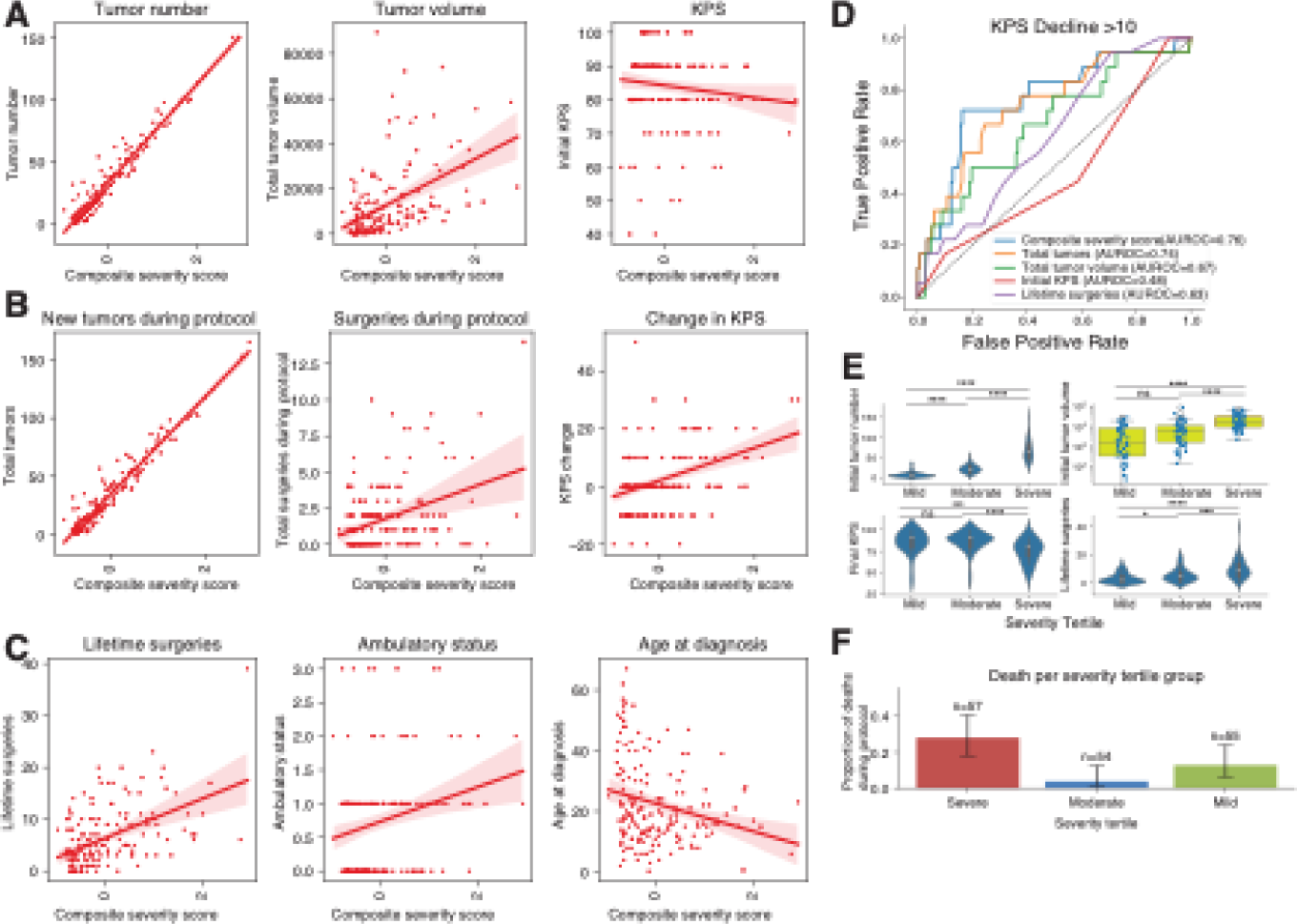
Composite severity score captures NF2-SWN disease burden and correlates with prospective clinical progression. **A.** Scatter plots demonstrating the relationship between the Composite enet base severity score and its baseline input parameters at study entry: total tumor count, total tumor volume, and Karnofsky Performance Status (KPS). Linear regression model fit lines with confidence intervals (95%) illustrate concordance between the composite score and individual components of baseline disease burden. **B.** Scatter plots of Composite enet base severity score with independent markers of prospective progression, including the number of new tumors that developed during the protocol, the number of tumor surgeries performed during the protocol, and the change in KPS over the study period. These relationships reflect the supervised tuning of the composite score to a predefined Progression Index. **C.** Scatter plots showing the relationship between the Composite enet base severity score and additional clinical variables not used in the score’s construction: lifetime surgeries, ambulatory status, and age at diagnosis. These associations demonstrate external face validity across complementary clinical dimensions. **D.** Receiver operating characteristic (ROC) curves evaluating the performance of the Composite enet base severity score and comparator clinical features for predicting a ≥10-point decline in KPS during the protocol. Area under the ROC (AUROC) values are shown for each parameter. **E.** Violin plots comparing composite score-defined tertiles (Mild, Moderate, Severe) against key clinical variables-including baseline tumor count, baseline tumor volume, KPS at study termination, and lifetime surgeries; demonstrating expected ordinal trends in disease burden across severity strata. Tukey post-hoc: ****: P<0.001; ****: P<0.00001. **F.** Bar plots depicting deaths (n=25) observed during the protocol with most deaths occurring in the severe tertile (χ² statistic = 11.36; P = 0.003).

**Fig. 4.**
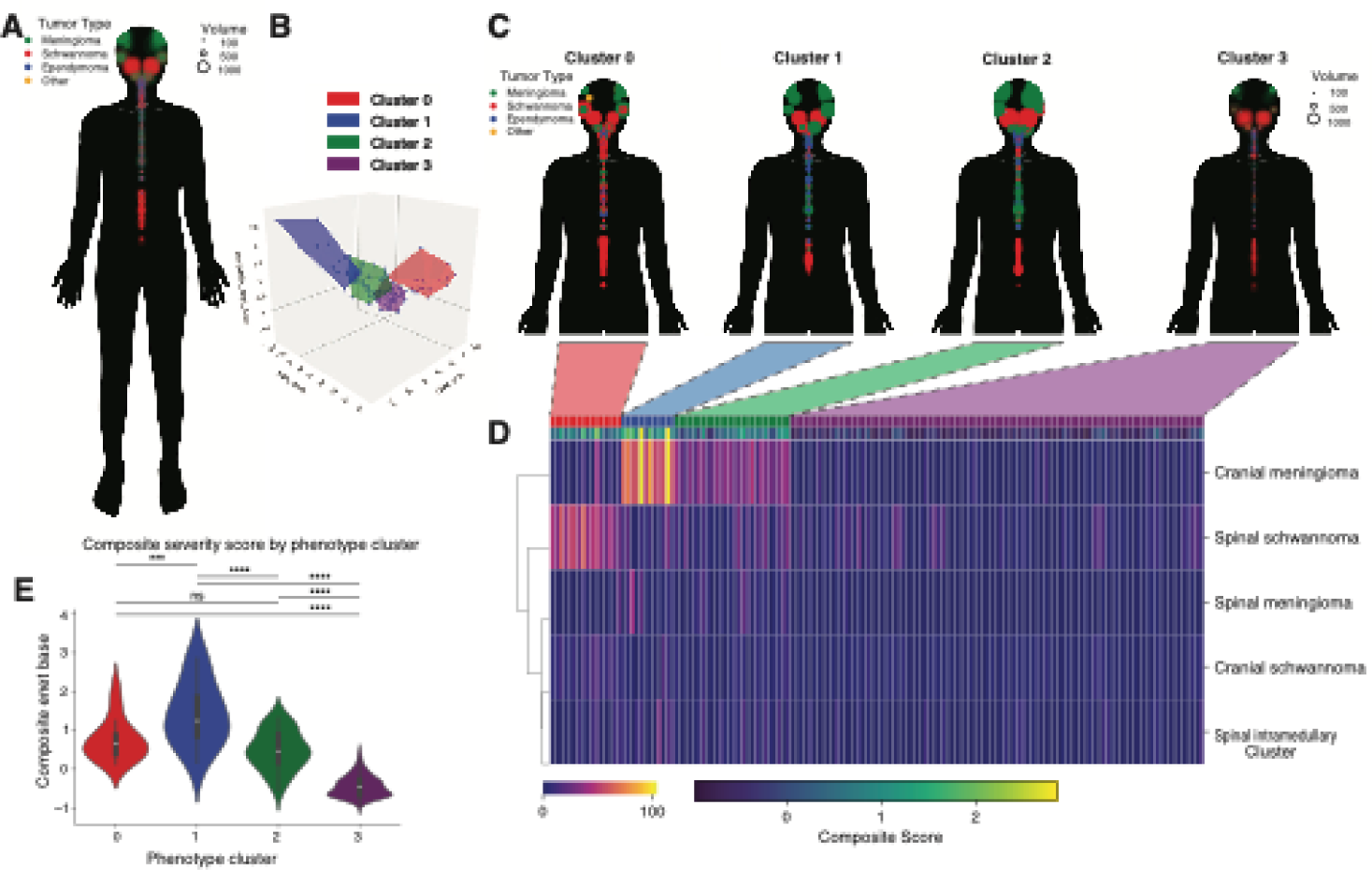
NF2-SWN patient phenotype clusters correlate with composite disease severity and tumor anatomic distribution. **A.** Anatomic map of all tumors (n = 5764) included in the cohort, plotted onto a generic human silhouette. Each point represents an individual tumor, with circle size proportional to tumor volume (mm³) and color indicating tumor type (meningioma, schwannoma, ependymoma, or other). This visualization highlights the wide distribution of cranial and spinal tumors in NF2-SWN. **B.** Principal component analysis (PCA) of NF2-SWN patient phenotypes using tumor burden metrics. Each point represents a patient plotted along PCA1, PCA2, and Composite severity score, with color shading the emergent clusters. Convex hulls are drawn around clusters in PCA space to illustrate separation of patient subgroups according to tumor burden and Composite severity score. **C.** Cluster-specific anatomic phenotype maps, showing spatial distribution of tumor types and tumor volumes for patients within each PCA-derived cluster (Cluster 0–3). Tumor locations, types, and sizes are overlaid on the same reference silhouette as in Panel A, revealing distinct anatomic patterns across phenotype clusters. **D.** Clustered heatmap summarizing per-patient tumor characteristics. Each column represents a patient and rows include cluster assignment, Composite enet base severity score, and counts of tumor subtypes (cranial meningioma, cranial schwannoma, spinal meningioma, spinal schwannoma, and spinal intramedullary tumors). **E.** Violin plots comparing Composite enet base scores across phenotype clusters. Tukey post-hoc: ****: P<0.001; ****: P<0.00001.

**Fig. 5:**
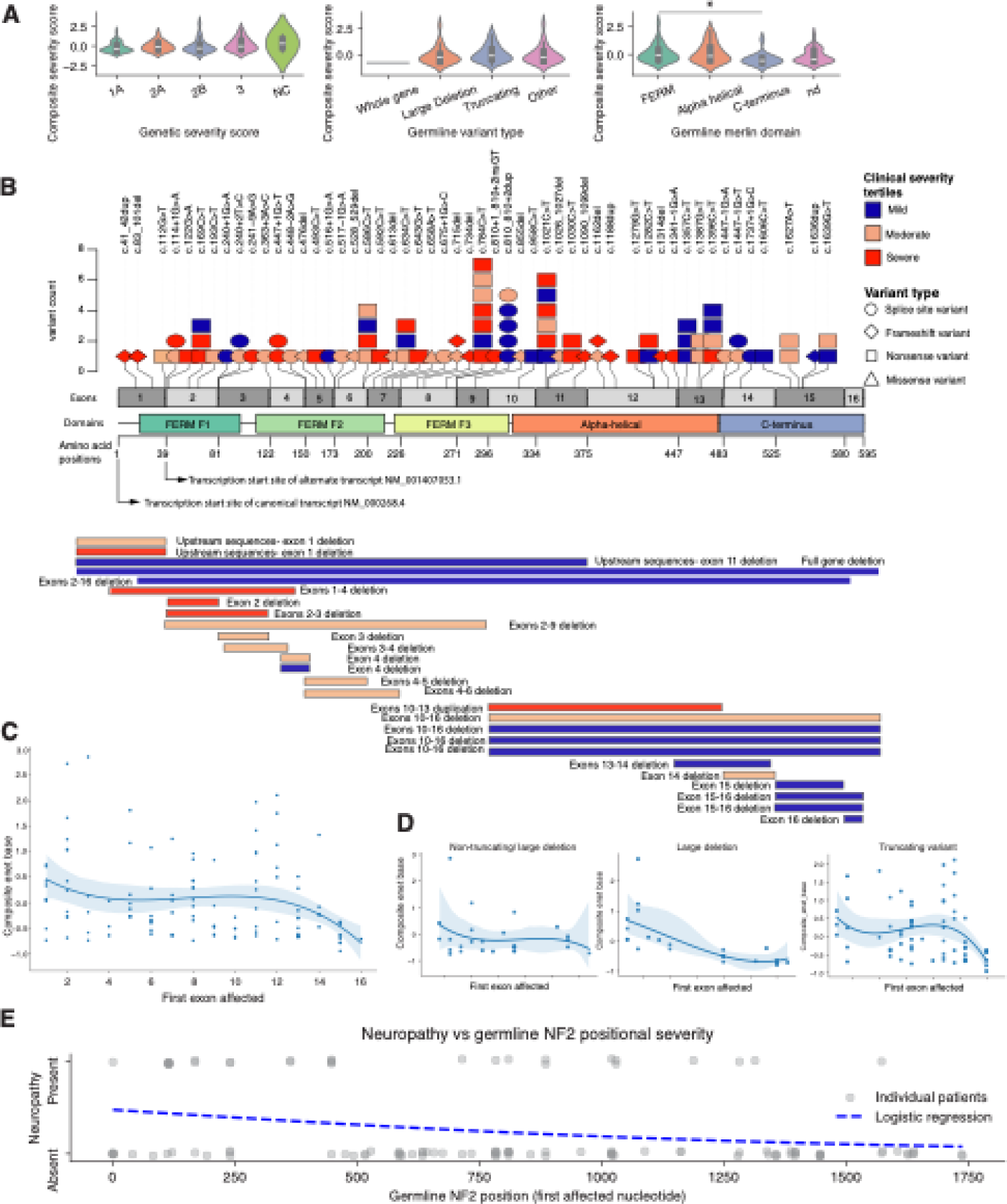
Location of the germline mutation affects disease severity. **A.** Parallel violin plots demonstrating the relationship of the composite severity score to genetic severity score, germline *NF2* variant type, and the location of germline variant on the *NF2* locus functional domains. Tukey post-hoc: *: P<0.05, ****: P<0.001; ****: P<0.00001. **B.** A lollipop plot mapping the location of the germline pathogenic single nucleotide variants and indels on the *NF2* locus with colors indicating the severity and shape indicating the type of variant. Exonic boundaries and functional Merlin domains are displayed for reference. Horizontal bars represent the genomic extent of copy number variants (deletions and duplications) within the *NF2* gene, colored by severity **C.** A scatterplot of germline variants on the *NF2* locus (x-axis), and Composite severity score (y-axis). The third-order polynomial regression line and 95% confidence interval is shown. **D**. Scatterplots showing the distribution of Composite severity score against each type of germline variant. **E.** Logistic regression plot depicting germline *NF2* variant position on the x-axis and presence of neuropathy on the y-axis (present/absent).

**Fig. 6.**
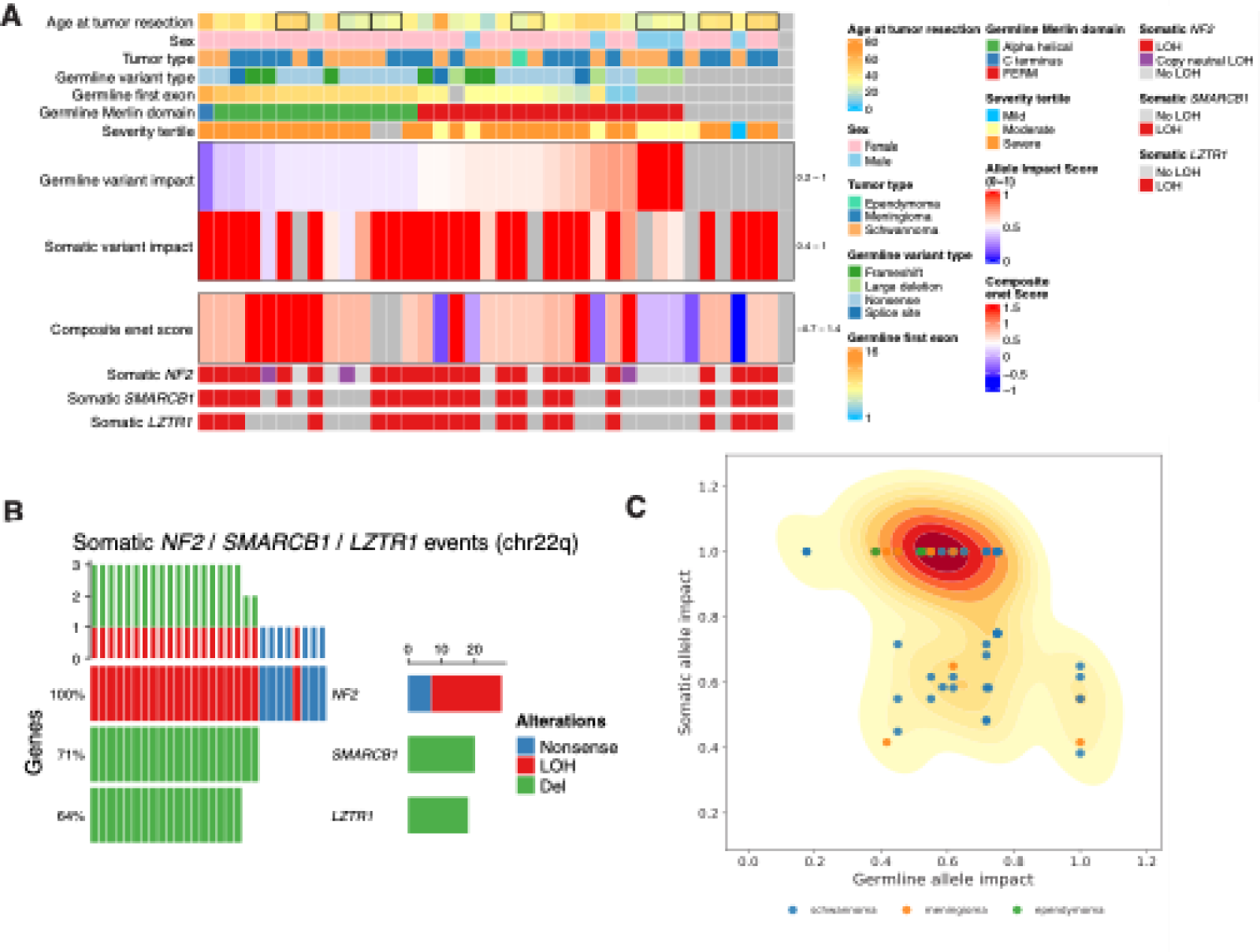
Germline pathogenic *NF2* variants constrain the landscape of second-hits. **A.** A heatmap of tumor specimens sequenced with the targeted gene panel (see Methods, and Fig. 1). Each column represents a single tumor, ordered according to the first germline nucleotide affected by a pathogenic variant. The top annotation rows summarize the phenotypic characteristics of the subjects. Tumors from same subjects are bound by boxes. **B.** An oncoprint plot assessing the frequency of somatic loss of chromosome 22q as the mechanisms of *NF2* loss of function. Top annotation summarized the additive effect of gene deletions. **C.** Two-dimensional kernel density estimate of germline allele impact (x-axis) versus somatic allele impact (y-axis) across all analyzed tumors (n = 77), with individual tumors overlaid as points colored by tumor type.

**Fig. 7.**
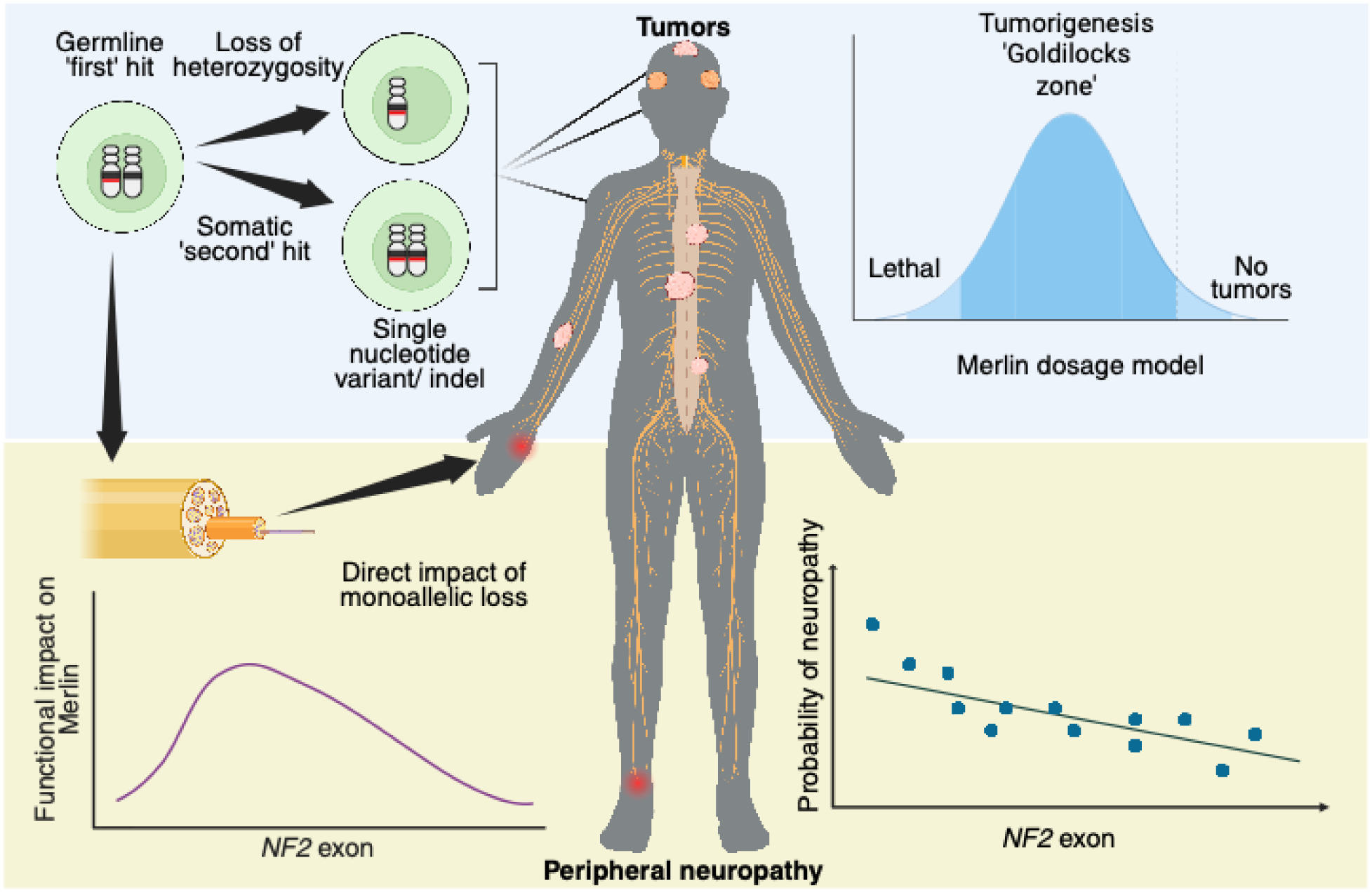
Dual mechanistic models of NF2-SWN: Merlin dosage-dependent tumorigenesis and variant location-dependent peripheral neuropathy. Tumor development in patients with germline *NF2* variants requires a second somatic hit that reduces merlin activity to a critical threshold - a “Goldilocks” zone-sufficient for tumorigenesis but not cellular lethality. By contrast, peripheral neuropathy in NF2-SWN arises through haploinsufficiency, driven solely by the location of the germline *NF2* variant.

### Multidimensional clinical and imaging features reveal broad phenotypic variability in NF2-SWN patients

NF2-SWN is clinically heterogeneous, but the extent and structure of variability in tumor burden, anatomic distribution, and functional outcomes have not been comprehensively defined. We first quantified phenotypic diversity by jointly analyzing tumor count, tumor volume, tumor location from whole-neuroaxis imaging, and functional status. Across the cohort, total tumor count and total volume varied by more than two orders of magnitude (**Fig. 2A**). Some patients with many tumors had modest total volume, whereas others with few tumors had very large lesions, indicating that different burden metrics capture distinct biological dimensions. Functional status (KPS) also varied widely and showed no simple relationship with tumor number or volume, underscoring the limitations of any single metric for capturing overall NF2-SWN severity.

We next cataloged phenotypic patterns (**Fig. 2B**) using longitudinal volumetric neuro-axis MRI. Even patients with similar tumor counts showed markedly different tumor distributions, and quantitative mapping of cranial versus spinal lesions revealed divergent anatomic patterns (**Fig. 2C**). We then integrated multiple disease dimensions-total tumor count, total volume, percent volume change over time, lifetime surgeries, and functional impairment (100 – KPS) into summary phenotypic plots. These multidimensional summaries (**Fig. 2D**) revealed distinct, non-overlapping phenotypic “shapes” across patients, highlighting substantial inter-patient variability. These findings led us to pursue a multi-dimensional approach to quantifying disease severity in NF2-SWN.

### GSS does not correlate with phenotypic variability or disease severity markers in NF2-SWN

We performed targeted deep sequencing of *NF2* and other schwannomatosis-and meningiomatosis-associated genes. About one-third of patients showed mosaicism (59/166, 35.5%), with *NF2* variants present at low allele fraction (<30%) or undetectable in peripheral blood. Most cases were de novo (111/166, 66.8%), lacking a documented family history of NF2-SWN. Pathogenic or likely pathogenic *NF2* variants included truncating (42%), splice-site (18%), and large deletions (16%) distributed across the gene (**Sup. Table 1**).

The Genetic Severity Score (GSS)(16) predicts NF2-SWN severity from germline genotype (variant type, location, mosaic status) and inferred functional impact, and has been replicated with moderate success in independent cohorts (11,14). However, GSS classes did not show monotonic or discriminatory relationships with traditional quantitative disease measures in our cohort (**Fig. 2E**). Thus, GSS could not account for the multidimensional phenotypic heterogeneity in our cohort, indicating a need to revisit genotype-based prediction strategy. We first attempted to capture the multidimensional nature of disease severity in NF2-SWN.

### Composite severity score captures NF2-SWN disease burden

To quantify baseline disease severity in NF2-SWN and enable forward-looking clinical stratification, we developed a multivariate Composite Severity Score (CSS) using only variables available at study entry. We selected five clinically relevant, moderately collinear baseline features (variance inflation factor range 2.6 - 4.6: total tumor count, total tumor volume, functional status by KPS, number of surgeries prior to protocol enrollment, and age at protocol enrollment (**Fig. 2A, Sup. Fig. 1A-C**). These were used to construct a baseline severity index. Unlike prior efforts, we intentionally excluded hearing loss, since we found that vestibular schwannoma-related hearing loss did not correlate with tumor size(18,19) or growth rate (20,21).

We next generated a Progression Index (PI) incorporating only prospective endpoints: number of new tumors arising during protocol, the number of surgeries during protocol, and change in KPS between enrollment and final visit. This design ensured that the supervised statistical model learned the links between baseline phenotype and subsequent clinical deterioration. We evaluated several strategies, including equal-weight averaging of baseline predictors, PCA, and supervised iterative regression using scikit-learn’s ElasticNetCV and LassoCV algorithms(22). Equal-weight and PCA approaches demonstrated moderate associations with the Progression Index (r ≈ 0.31–0.36) and fair predictive capacity for KPS decline ≥10 points (AUROC 0.63–0.65). PCA was not favored since its loadings showed limited translational applicability as a prognostic tool.

In contrast, a supervised Elastic Net model trained on the same baseline variables (Z scored tumor number, total tumor volume and KPS) achieved the best performance. This model provides penalized feature selection to reduce overfitting and stability in small sample sizes, yielding a reproducible, formula-based severity score we call the composite severity score (CSS). This approach produced a parsimonious set of variables (**Sup. Fig. 1A**) that contributed to the CSS – initial tumor count and initial KPS. The CSS distribution mirrored the input parameters (**Sup. Fig. 1B**).

The CSS correlated strongly with its baseline inputs (**Fig. 3A**) and was significantly associated with the prospective progression index (**Fig. 3B**), despite being constructed exclusively from retrospective data. It also correlated with independent markers not used in its construction including lifetime surgeries, ambulatory status, and age at diagnosis (**Fig. 3C**). CSS values showed a monotonic relationship with stepwise KPS decline during follow-up (**Sup. Fig. 1C**), supporting its use as a clinically relevant predictive tool.

Using bootstrapping for internal validation, the CSS correlated with the Progression Index at r=0.54, with minimal optimism, indicating the relationship is not driven by overfitting. Tumor count was the dominant, stable predictor (weight=0.77, 95% CI: 0.64–1.00); KPS contributed a smaller, consistent effect (weight=−0.23, 95% CI: −0.36–0.00).

### Composite severity score correlates with prospective clinical progression

We assessed predictive discrimination of the CSS using AUROC across several clinically relevant outcomes. The CSS displayed the highest AUROC (0.76) among all evaluated predictors for identifying patients who experienced a ≥ 10-point KPS decline during the study protocol (**Fig. 3D**). It also demonstrated fair discrimination for new tumor onset (0.67), protocol surgeries (0.63), lifetime surgeries (0.73), and ambulatory impairment (0.6). Stratifying patients into CSS tertiles (mild, moderate, severe) showed clear ordinal separation across multiple retrospective and prospective disease measures including baseline tumor count, baseline mass burden, end-of-study KPS, and lifetime surgeries (**Fig. 3E**, Tukey post-hoc, P < 0.001–0.00001). We recorded 25 deaths during the protocol (**Fig. 3F**), a majority of whom (n = 14) were in the severe CSS tertile (χ² statistic = 11.36; P = 0.003). These findings suggest that CSS score is a multidimensional measurement of baseline heterogeneity that meaningfully predicts clinical progression.

### NF2-SWN phenotypic clusters correlate with disease severity

To determine whether NF2-SWN patients exhibit phenotypic subgroups based on their anatomic tumor distribution, we performed unsupervised clustering using scaled counts of five tumor classes across the neuroaxis (cranial meningiomas, cranial schwannomas, spinal meningiomas, spinal schwannomas and intramedullary tumors).

We coded the anatomic location of each of the 5,764 tumors measured in the study to generate an anatomic distribution on a generic human silhouette (**Fig. 4A**, **Sup. Fig. 2A**). The map captured the distinct craniospinal distribution each tumor type (**Sup. Fig. 2A**). When projected onto PCA space, patients formed four discrete tumor clusters, each occupying a distinct region of the PCA1-PCA2 plane and demonstrating separation along the CSS gradient (**Fig. 4B**).

Cluster-specific anatomic maps demonstrated distinct spatial signatures. Clusters 1 and 2 were enriched for meningiomas. Spinal schwannomas were more frequent in cluster 0, while spinal meningiomas predominated in cluster 1 (**Fig. 4C and 4D**). Quantitatively, CSS values varied inversely with tumor clusters. Clusters showed well-separated severity distributions (**Fig. 4E, Sup. Fig. 2B**); cluster 3 exhibited predominantly mild CSS values, whereas clusters 0, 1, and 2 were enriched for severe cases (χ² = 110.8, p < 0.0001). These findings indicate that phenotypic clustering captures meaningful variation in disease burden, and that CSS tracks cluster membership.

We asked whether these phenotype-defined tumor clusters corresponded to *NF2* genotype categories, including truncating variants, splice-site variants, large deletions, missense variants, or classical genotype-severity frameworks such as the GSS, mosaicism, or merlin domain involvement. No statistically significant associations emerged between genotype and cluster membership across all comparisons including truncating/non-truncating variant types, GSS, mosaicism, and domain-level annotations (FERM, α-helical, C-terminus; **Sup. Fig. 2C and 2D**; χ² p-values 0.127-0.729). Thus, we found that genotypic categorization alone did not determine the observed phenotypic clustering in our cohort.

However, when individual tumor types were examined across merlin structural domains, there was a significant association between C-terminal *NF2* variants and schwannoma burden. These patients carried significantly fewer schwannomas compared with patients harboring FERM domain variants (Tukey HSD p = 0.026; **Sup. Fig. 2E**). This raises the possibility that *NF2* variant position may modulate specific tumor phenotypes despite a lack of clear association with phenotypic clustering patterns.

### Impact of mosaicism and germline variant location on NF2-SWN disease severity

Previous studies indicated that *NF2* mosaicism is associated with milder disease and these individuals were classified into lower severity GSS categories(12,23), although mosaicism can also be linked to markedly variable phenotypes, including increased clinical severity is some patients (11). In our cohort, mosaic status did not correlate with multiple metrics of disease burden, including CSS (ANOVA P = 0.214; all Tukey P > 0.18), total tumor count (ANOVA P = 0.222; all Tukey P > 0.21), and baseline tumor volume (ANOVA P = 0.105; all Tukey P > 0.13; **Sup. Fig. 3**), indicating that mosaicism alone did not explain the wide phenotypic variability observed in our cohort.

Likewise, other genotypic variables such as variant type (GSS, nonsense, frameshift, splice-site, large deletion) did not show consistent or monotonic relationships with clinical severity (**Fig. 5A**). However, when we mapped each germline variant to its location along the *NF2* locus and corresponding merlin domain, we observed that rather than a smooth decline in severity from F1 to C-terminus (24,25), patients with germline variants within the early F1 and the α-helical domains exhibited elevated disease severity, illustrated using a third-order polynomial regression plot (**Fig. 5B-D**), while F2, F3 and C-terminus variants conferred less severity. Following an initial dip, the severity curve showed a modest distal resurgence in the α-helical region, suggesting that merlin structure-function constraints, rather than residual protein length, affect the clinical phenotype. This relationship was observed across several severity metrics including total tumor count, total mass burden, and final KPS (**Sup. Fig. 4**). Together, these findings indicate that germline variant location is a more informative predictor of disease severity than variant type.

### Neuropathy patterns support a germline merlin dosage effect

Since NF2-SWN-associated neuropathy is driven primarily by merlin haploinsufficiency(9,26,27) rather than biallelic loss, we examined whether neuropathy prevalence reflected the positional effects observed for tumor severity. Neuropathy was more common among individuals with FERM-domain germline variants and least frequent in those with C-terminal variants, a trend visible in domain-level proportions and supported by modeling of neuropathy incidence with germline position (**Fig. 5E**). Logistic regression demonstrated that neuropathy risk increased significantly toward the 5′ end of *NF2* (β = –0.0014, p = 0.004). These findings suggest that the germline *NF2* allele alone can exert a measurable biological effect on neurons, modulated by the location of the variant.

### Germline NF2 pathogenic variants constrain the landscape of somatic second hits

We performed targeted deep sequencing of 37 surgically resected, symptomatic tumors (NCT00060541) to characterize the somatic mutational landscape of NF2-SWN across the full range of clinical severity, germline variant classes, merlin domain groups, and patient sex, including multiple tumors from some individuals (**Fig. 6A**). The dominant somatic mechanism across tumors was chromosome 22q LOH, evidenced by frequent co-deletion of *NF2*, *LZTR1*, and *SMARCB1* (detected in 60% of tumors; **Fig. 6B, Sup. Table 2**). Tumors from this sequencing cohort with complete germline and somatic *NF2* hit information were combined with tumors identified from the published literature (6,28,29) meeting the same criterion, yielding a final cohort of 77 tumors from 63 patients **(Sup. Methods 1.7)**.

To test the relationship between the germline and somatic events, variants were assigned continuous allele impact scores (0–1 scale) based on predicted merlin loss. Whole-gene deletion and loss of heterozygosity (LOH) were modeled as maximally severe, while and other variants modeled as producing intermediate merlin activity **(Sup. Methods 1.8).** Visualizing the joint distribution of germline and somatic allele impact across all 77 tumors revealed non-random structure rather than a uniform spread of combinations. Three tumor groups were identified: a predominant cluster (n=51) characterized by moderate germline impact paired with maximal somatic impact (somatic LOH), an intermediate cluster (n=20) with moderate impact on both alleles, and a smaller cluster (n=6) combining maximal germline impact with comparatively lower somatic impact (**Fig. 6C**). Severe-severe and mild-mild allele combinations were conspicuously absent. The clustering structure was significantly stronger than expected by chance (silhouette coefficient = 0.62; permutation p = 0.003, 1,000 iterations). These findings align with previous studies (25,27) that complete loss of merlin function is incompatible with cell survival, and we further propose that insufficient loss is likely biologically inconsequential for tumor formation. Thus, In the setting of a severe germline allele, a high impact somatic event would be predicted to induce cell death rather than tumorigenesis, effectively lowering disease severity in these individuals. In line with our hypothesis, all 3 patients in our cohort carrying a germline whole/near-whole gene deletion were classified as mild phenotypes by CSS.

We also observed that meningiomas were significantly enriched in the predominant moderate-germline/maximal-somatic-LOH cluster relative to the other two clusters combined (odds ratio = 4.17, Fisher’s exact p = 0.021), suggesting that meningiomas arising in NF2-SWN may preferentially develop via somatic LOH as the second hit. While the small size of some subgroups limits definitive conclusions, this pattern is consistent with the broader hypothesis that specific combinations of germline and somatic allele impact — rather than germline or somatic severity independently — shape tumor phenotype in NF2-SWN.

## Discussion

NF2-SWN is a devastating neurological disorder associated with severe morbidity and early mortality, driven by widespread central and peripheral nervous system manifestations, and due to postoperative complications (4,31). Previously, missense and splice-site variants, large deletions and mosaicism have been associated with milder phenotypes, and truncating variants to more severe disease (12,32,33). However, in our cohort, deep phenotyping and targeted NGS did not reveal consistent associations between clinical severity and variant type (except milder disease with germline large deletion). Importantly, mosaicism was not associated with milder disease manifestation, suggesting that other variables were responsible for the final phenotype.

To address limitations of prior genotype-phenotype studies, we constructed a multivariate Composite Severity Score (CSS) from clinically measurable variables at study entry. We then derived a longitudinal Progression Index (PI) capturing new tumors, surgeries, and KPS change, and used an Elastic Net model (composite severity score) to fit PI to CSS and refine the CSS weights. The final CSS derivation was optimized with a parsimnonious set of input variables – total tumor number, and KPS at study entry. The CSS then correlated with age at diagnosis, predicted new tumor onset, protocol and lifetime surgeries, ambulatory impairment, and importantly, KPS decline during the study duration. The CSS achieved the highest AUROC (0.76) for identifying patients with a ≥10-point KPS decline among all predictors tested.

Previous work has examined the clinical impact of *NF2* variant position. Missense variants (∼5%) are too infrequent for robust genotype-phenotype correlations, and splice-site variants (∼25%) show variable severity, with changes in exons 1-5 associated with worse outcomes than those in exons 11-15 (34). Likewise, large deletions spanning exons 1-9 have been linked to earlier symptom onset (19 vs 30 years for deletions in exons 10-15)(34), and meningiomas are rarely seen in patients or tumors with splice-site or somatic *NF2* variants in exons 14-15 (35). In our study, germline variants in the early F1 FERM domain (exon 1) or α-helical domain (exons 10-13) were associated with higher overall severity, and neuropathy was more frequent in patients with FERM-domain variants, whereas C-terminal variants were associated with significantly fewer schwannomas.

Post-translational modifications are one of several mechanisms that confound NF2-SWN genotype-phenotype associations and contribute to the non-linear relationship between *NF2* expression and merlin protein levels (5). Other confounders include epigenetic changes such as DNA methylation and *NF2*-associated miRNAs (36). Downstream signaling pathways likewise influence disease severity. In a parallel paper (8), we identified a role for lineage-specific downstream *TEAD1* expression in schwannomatosis in NF2-SWN patients, and inhibition of YAP/TAZ-driven TEAD signaling shows therapeutic potential in *NF2*-null mouse cells (37). Peripheral neuropathy also contributes to NF2-SWN disease severity (2,9). Unlike Schwann cells, terminally differentiated neurons do not replicate and have fewer opportunities to acquire somatic second hits (6). Thus, NF2-SWN neuropathy reflects germline haploinsufficiency, with neurons transitioning from normal axonal function at near-diploid merlin dosage to neuropathy as merlin levels approach haploidy.

In the tumor-suppressor gene *APC*, prior work has shown that the nature of the first hit shapes the nature of the second hit (38). To our knowledge, this concept that germline *NF2* variants constrain allowable second-hit somatic variants capable of driving tumorigenesis has not been previously applied to NF2-SWN. In humans, a second hit on the normal allele, and inactivation or loss of both alleles is seen in NF2-associated and sporadic schwannomas and meningiomas (39,40). In mice however, heterozygosity is insufficient to inactivate the second allele likely due to the relative rarity of a second hit, and heterozygous mice mainly develop osteosarcomas whereas schwannomatosis requires conditional homozygous *Nf2* knockout (41,42)(52–55). We propose that in patients with severe germline variants, a severe somatic second hit is likely cytotoxic, and tumor formation depends on a “Goldilocks” combination: either two moderate variants (one germline, one somatic) or one severe (germline or somatic) plus one moderate variant.

Although early truncating F1 variants would be expected to behave as high-impact alleles, an in-frame methionine in exon 2 and a downstream transcription start site likely permit partial rescue of merlin expression, so these variants behave as moderate impact variants unless combined with LOH. In contrast, we identified 3 tumors with full or near-full gene deletions which, when combined with non-LOH second hits, resulted in tumorigenesis. Other studies report 14 additional patients with germline full/near-full deletions and tumors carrying non-LOH second hits (6,7,29,43,44).

Additionally, across 77 tumors we identified specific combinations of germline and somatic alleles associated with tumorigenesis, with a notable absence of biallelic mild-mild or severe-severe combinations.Together, these data suggest that the first hit (germline variant) constrains the second hit (somatic alteration). Thus, we hypothesize that the somatic landscape is non-random and bounded: tumors develop only when germline and somatic hits together reduce merlin activity to a tumor-permissive but non-lethal range, supporting a dosage-based model of merlin function (**Fig. 7**).

Our study is limited by single-institution data, modest sample size, inferred functional merlin levels based on variant location and limited follow up. Moreover, our germline-somatic interaction analysis does not account for multiple *NF2* somatic variants in some tumors or epigenetic silencing of the second allele. However, our consistent prospective collection of serial deep phenotypic data permitted analytic insights not previously reported. Future large prospective studies and graded merlin depletion experiments are needed to replicate our key findings.

## Conclusions

In this study, we used deep phenotyping from a prospective single-institution database to develop the CSS for NF2-SWN that integrates disease burden (tumor count), and functional impact (KPS). CSS outperformed existing single variables and predictive models for estimating NF2-SWN severity and enabled identification of patient clusters that grouped by severity. Tumor analysis revealed a limited set of germline-somatic variant combinations, suggesting that the genomic location of the germline variant constrains permissible second-hit somatic mutations. These findings provide a foundation for future therapeutic strategies for NF2-SWN.

## Data Availability

Sequencing datasets generated in this study are not publicly available due to ethical and privacy considerations related to patient confidentiality; however they can be made available upon reasonable request and subject to institutional and ethical approvals. All other data generated or analyzed as part of this research have been included in the manuscript and accompanying supplementary data.

## Acknowledgements

The authors are clinicians dedicated to patient care. The authors thank the patients and their families for participating in the clinical trials that made this work possible.

## Supplemental Material

### Supplemental Methods

#### 1.1 Study design

This is a prospective, longitudinal cohort study of patients meeting the clinical criteria for NF2-SWN and/or genetic testing demonstrating pathogenic germline NF2 variants, or identical variants in the NF2 gene in tumors from two separate sites. Other inclusion criteria included age at enrollment between 8 and 75 years, ability to undergo serial magnetic resonance imaging (MRI) scans with gadolinium-based contrast and capacity to provide consent (or the availability of a legal guardian to provide consent in certain circumstances). Exclusion criteria included clinical comorbidities that precluded clinical evaluation and serial imaging, such as severe renal insufficiency, hepatorenal syndrome, history of liver transplantation, and pregnancy at the time of visit. Serum and peripheral blood mononuclear cells (PBMCs) were collected during each visit. The study was approved by the Combined Neuroscience Institutional Review Board of the National Institutes of Health (IRB approval: 08-N-0044, NCT00598351). Written and informed consent was obtained from every participant prior to enrollment. The study was conducted in accordance with the principles of the Declaration of Helsinki.

#### 1.2 Clinical evaluation

High resolution MR imaging (1–1.5 mm slices) with gadolinium was performed. Patients with measurable hearing underwent auditory evaluation, while vestibular testing was performed in those without prior treatment for vestibular schwannoma. Additional testing included speech/swallow exams, neurophysiology, and imaging as clinically indicated. At clinic visits, patients or caregivers completed surveys (Speech and Swallow Questionnaire, Functional Independence Measure). Clinicians assessed Karnofsky performance status (KPS), ambulatory status (Modified McCormick Scale), and abbreviated ASIA motor scores. Yearly assessments included audiograms, vestibular testing (if untreated or symptomatic), and speech/swallow or neurophysiology as indicated. Non-serviceable hearing was defined as speech recognition <50% and/or PTA >50 decibels. MRI of the brain and spine with/without contrast was performed at each visit, with additional imaging as needed. Patients were followed annually or sooner for new/worsening symptoms. If follow-up was not feasible, questionnaires were mailed (when possible), and outside imaging was obtained for review. Patients were followed for up to 5 years, with continued care for surgery-related complications. Surgery for NF2-SWN-related conditions was performed per standard of care.

#### 1.3 Data collection and image analysis

Patient data was obtained via electronic medical record review. MRI scans performed at the time of enrollment and the most recent follow up were compared. Tumors were identified by imaging characteristics if pathology was unavailable. Tumor volumes were measured manually by a single investigator for each patient using the ABC/2 method. Additional features included diffuse meningeal thickening (meningiomatosis), tumor extension into the face via skull base foramina or bone invasion, and innumerable punctate cauda equina lesions. Tumor burden was calculated from total tumor volume and number.

#### 1.4 Genetic Testing

Genetic testing from CLIA-certified laboratories was available in 69 patients. In-house genetic testing was performed on 125 patients using a custom amplicon-based targeted panel generated according to previously published methods (1) including all exons as well as flanking segments of the *NF2* gene as well as five other genes with potential overlapping phenotypes: *SMARCE1*, *LZTR1*, *SMARCB1*, and *SUFU*. The 5’ untranslated region and promoter of NF2 were also included.

DNA was extracted and purified from frozen PBMC pellets and tumors embedded in Optimal Cutting Temperature compound using the Qiagen AllPrep DNA/RNA extraction kit (Catalog no: 80204, Qiagen, USA). Purified DNA was quantified using the Qubit 4 fluorometer (ThermoFisher Scientific, USA). Libraries were prepared using the AmpliSeq custom DNA panel for Illumina, AmpliSeq Library Plus and AmpliSeq CD indexes for Illumina (Illumina, USA) according to manufacturer’s instructions. Sequencing was carried out on the Illumina MiSeq sequencer (Illumina, USA). Germline sequencing data was processed using Qiagen Clinical Insight Interpret (version 9.0.0.20220826) to identify sequence and copy number variants. Further processing for mosaic variants with allele frequency cutoffs <30% was carried out on Qiagen CLC Genomics Workbench 22.0.2 (https://digitalinsights.qiagen.com/). All variants were classified according to the American College of Medical Genetics/Association of Molecular Pathologists guidelines (2). Tumor sequencing data was processed on the NIH Biowulf high performance computing cluster using the Sarek nf-core pipeline (3). Specifically, somatic variant calling and annotation were performed using Strelka2 (4) and SnpEff (5) respectively. Copy number/ loss of heterozygosity analysis was carried out using cnvkit (6) and Manta (7) using default parameters.

#### 1.5 Construction and Validation of the Composite NF2-SWN Severity Score Baseline Feature Selection and Preprocessing

To construct a retrospective baseline severity metric applicable at the time of study enrollment, we selected three clinically relevant and minimally collinear variables reflecting NF2-SWN disease burden at baseline: total tumor count, total tumor volume, and KPS. Tumor count and tumor volume were extracted from whole-neuroaxis MRI annotations, while KPS was recorded at the baseline clinical encounter.

All variables were standardized using z-scoring based on cohort-wide means and standard deviations. KPS was inverted (Inverted KPS=−Z(KPS)) so that higher values consistently indicated worse disease severity. No post-enrollment variables were included at this stage to prevent information leakage.

### Forward-Looking Progression Index

To enable supervised model tuning, we generated a Progression Index composed exclusively of prospective clinical outcomes. This included number of new tumors that developed during protocol follow-up, number of tumor surgeries performed during the protocol period, and the change in KPS between study entry and final visit.

Each component was standardized, and the mean of these three z-scores was used as the composite Progression Index. This approach linked baseline disease features to subsequent clinical deterioration without incorporating any post-enrollment information into score construction.

### Evaluation of Candidate Composite Strategies

We compared multiple strategies for creating a multivariate severity index. Equal-weight composite comprised of the mean of the standardized baseline variables. Principal Component Analysis (PCA) where PC1 was used as an unsupervised severity axis. Penalized regression using supervised scikit-learn Elastic Net (8) and LASSO (9) algorithms that were tuned to the Progression Index. PCA captured variance structure but lacked outcome-directed weighting, limiting its utility for prognostic modeling. Equal-weight and PCA-based methods were retained as comparators.

### Elastic Net Model for Final Composite Score

The primary Composite Severity Score was derived using a supervised Elastic Net regression model implemented in scikit-learn (ElasticNetCV) (10) The model was trained using: 5-fold cross-validation, l1_ratio grid = [0.1, 0.3, 0.5, 0.7, 0.9, 1.0], random_state = 42 for reproducibility. Inputs were the three standardized baseline variables, and the target was the Progression Index. Elastic Net was selected because it provides (i) shrinkage to reduce overfitting, (ii) robustness in small-to-moderate sample sizes, and (iii) interpretable weights suitable for formula-based clinical application.

Coefficients were normalized by the sum of their absolute values to produce the final Composite_enet_base score: CSS=0.802*Z(Tumor Count) + 0.197*(-Z(KPS) with higher values indicating more severe disease.

### Prospective Validation of CSS

The Composite Severity Score was evaluated for correlation with baseline features (face validity). Next, the CSS was evaluated for correlation with individual prospective endpoints that define the Progression index. Lastly, associations with independent clinical markers: lifetime surgeries, ambulatory status, age at diagnosis were analyzed. Predictive discrimination using receiver operating characteristic (ROC) analysis was performed for ≥10-point KPS decline during protocol, occurrence of any new tumors, need for tumor surgery during protocol, lifetime tumor surgeries, and ambulatory impairment. Area under the ROC curve (AUROC) was calculated for the Composite Severity Score and comparator clinical variables.

### Internal validation of CSS

We assessed the two-predictor Composite Severity Score (standardized tumor count and inverted KPS) using bootstrap internal validation (1,000 resamples) to correct for overfitting, refitting the full Elastic Net pipeline on each resample. Optimism-corrected performance was calculated as the difference between bootstrap and original-sample fit. Because normalized coefficients distort raw R², we recalibrated by regressing the Progression Index on the composite score.

### Severity Stratification by Tertiles

To produce clinically interpretable severity groups, CSS values were stratified into tertiles (Mild, Moderate, Severe). Tertiles were chosen a priori to avoid overfitting of cut points and to ensure evenly sized groups suitable for downstream comparisons. Group differences across retrospective and prospective disease dimensions were assessed using ANOVA with Tukey post-hoc testing.

#### 1.6 Phenotypic clustering

Tumor phenotype clustering used five craniospinal burden metrics (cranial meningiomas, cranial schwannomas, spinal meningiomas, spinal schwannomas, spinal intramedullary tumors), excluding patients with missing data. Tumor counts were z scaled and grouped by k means clustering, with the optimal number of clusters (k = 1-10) chosen using the elbow method. Cluster assignments were visualized by principal component analysis (PCA) using PCA1-PCA2 plots with convex hulls outlining each cluster. For anatomical visualization, 5764 tumors were mapped onto a standardized human silhouette, where point size represented tumor volume and color indicated tumor type, and cluster specific maps were generated according to each patient’s cluster membership.

#### 1.7 Cohort assembly for germline-somatic variant analysis

Tumor-level germline and somatic *NF2* variant data were compiled from our institutional cohort together with cases reported in the published literature (1,11,12). Only tumors with complete information on both the germline (first) and somatic (second) *NF2* hits were included, consistent with the two-hit tumor suppressor model. Tumors harboring more than two identified *NF2* alterations, or unusual variant types not readily classifiable within our scoring framework (e.g., duplications), were excluded from analysis. This yielded a final analytic cohort of 77 tumors, including 63 patients from our institutional dataset combined with tumors identified from the literature.

#### 1.8 Allele impact scoring and analysis

To quantify the predicted functional severity of germline and somatic *NF2* variants, we developed an allele impact score (0–1 scale) incorporating variant type, exon position, and, where applicable, extent of genomic deletion. Whole-gene deletion and loss-of-heterozygosity (WGD/LOH) events were assigned the maximum score (1.0), reflecting complete loss of the allele. For single nucleotide variants (nonsense, frameshift, splice-site, and missense), severity was estimated using exon position as a proxy for the extent of functional protein retained. Exon position scores declined linearly from 0.75 (exon 2) to 0.35 (exon 14), reflecting progressively diminished predicted impact across the majority of the coding sequence. Exon 1 was assigned the midpoint of this range (0.55) to account for the possibility of translation reinitiation from a downstream alternative start site. Exons 15 and 16, located near the C-terminus, were assigned substantially reduced scores (0.175 and 0.035, respectively) to reflect a lower predicted impact near the gene terminus. For deletions, reading frame determined the scoring approach: out-of-frame deletions were scored using the same exon-position lookup as other truncating variants, while in-frame or frame-unknown deletions were scored proportionally to the percentage of the gene deleted, scaled to the range 0.5–0.95 (impact = 0.5 + [%deleted/100] × 0.45). Variants with missing or unclassifiable data were excluded from analysis.

To formally evaluate whether tumors segregated into distinct germline-somatic impact groups, we applied k-means clustering (k=3) to the paired (Germline_impact, Somatic_impact) values. Cluster quality was assessed using the silhouette coefficient. To determine whether the observed clustering structure exceeded what would be expected by chance—given that a subset of somatic scores are fixed at a ceiling value (1.0) for WGD/LOH events—we performed a permutation test in which somatic impact values were randomly shuffled relative to germline impact values (1,000 iterations), breaking any true germline-somatic pairing while preserving each variable’s marginal distribution. A permutation p-value was calculated as the proportion of shuffled datasets achieving a silhouette score equal to or greater than the observed value. Two-tailed Fisher’s exact test was applied to test if specific tumor types were over-represented in density plot clusters.

## Supplemental Tables

**Sup. Table 1:**
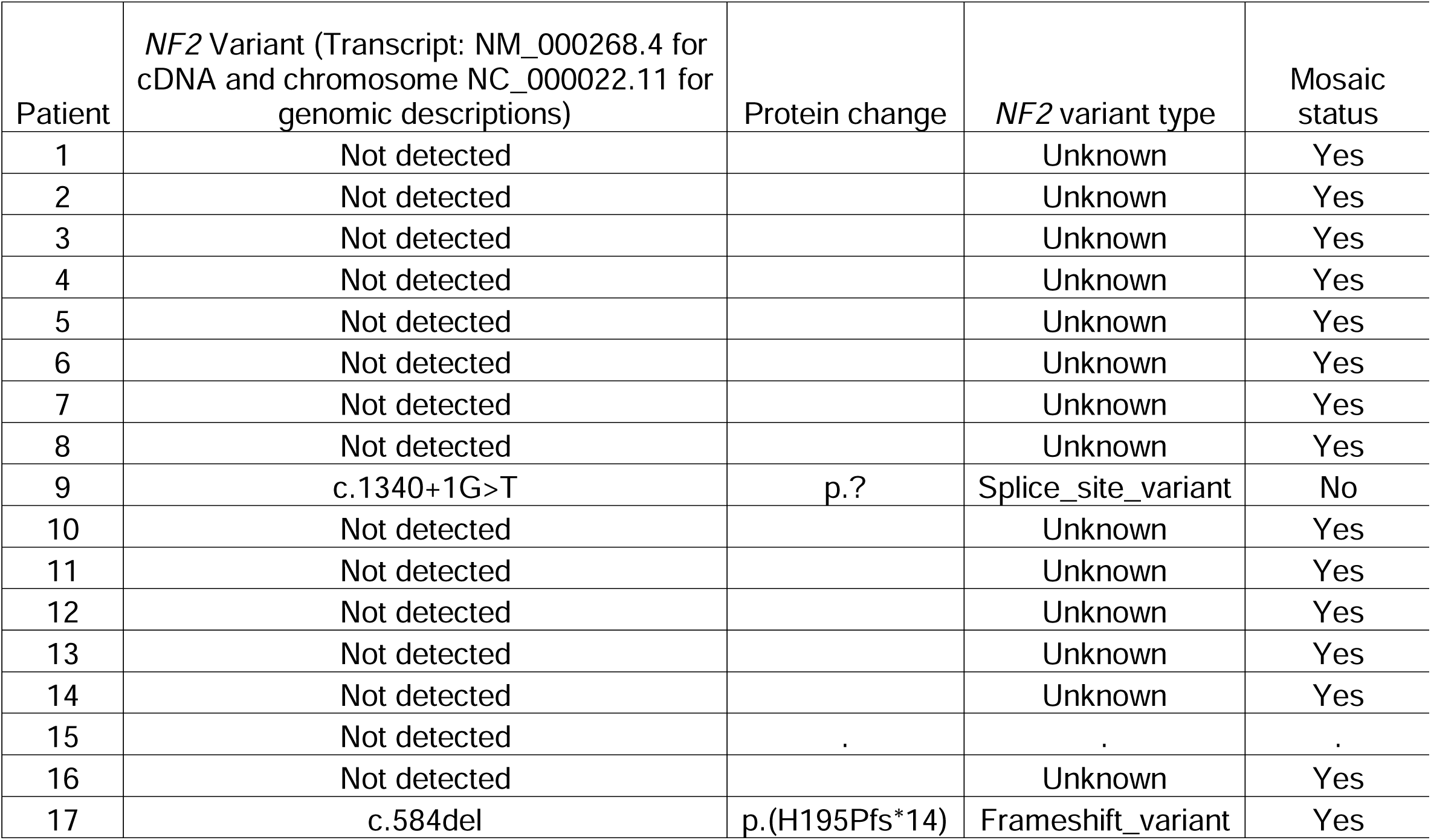
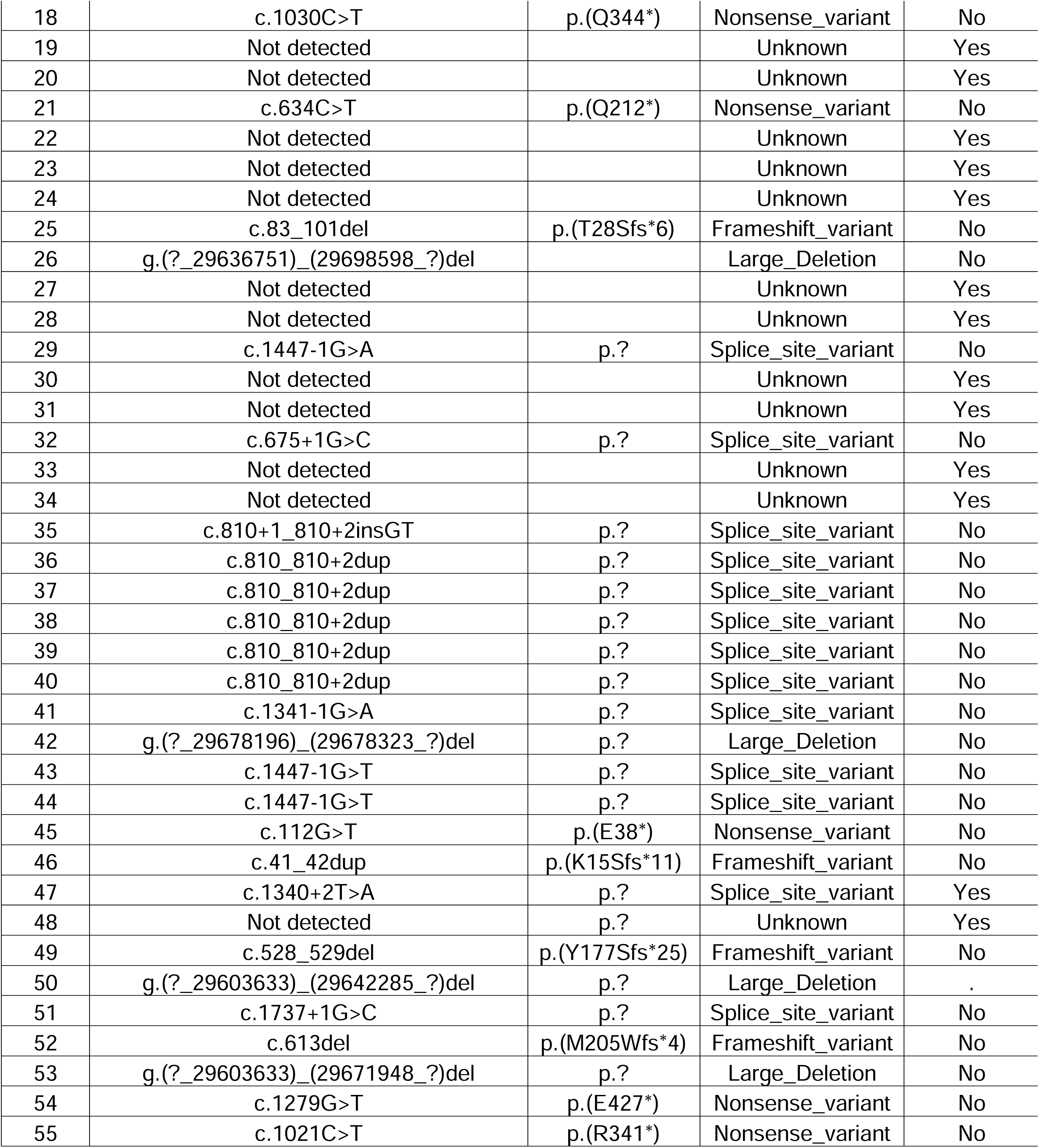
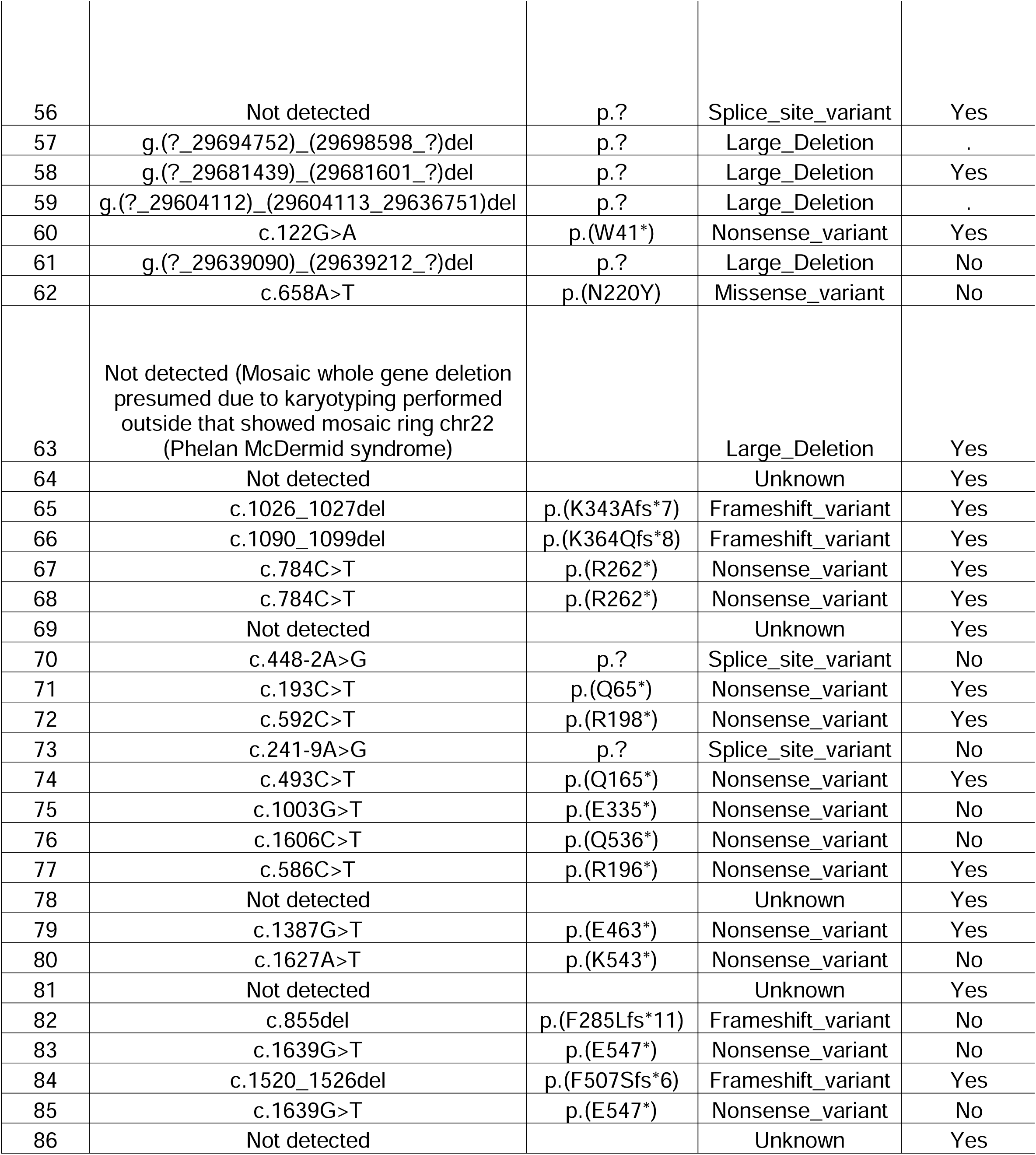
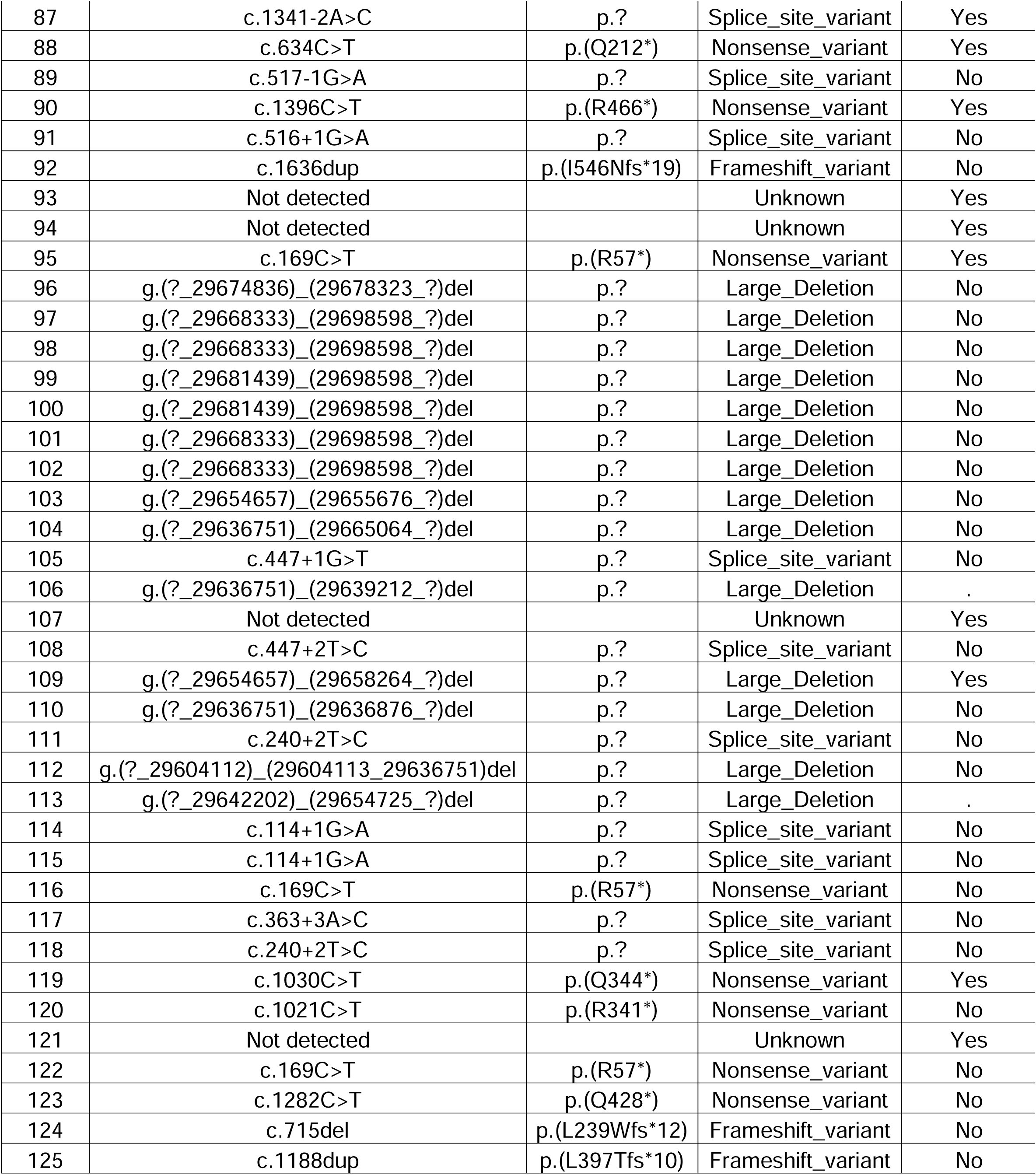
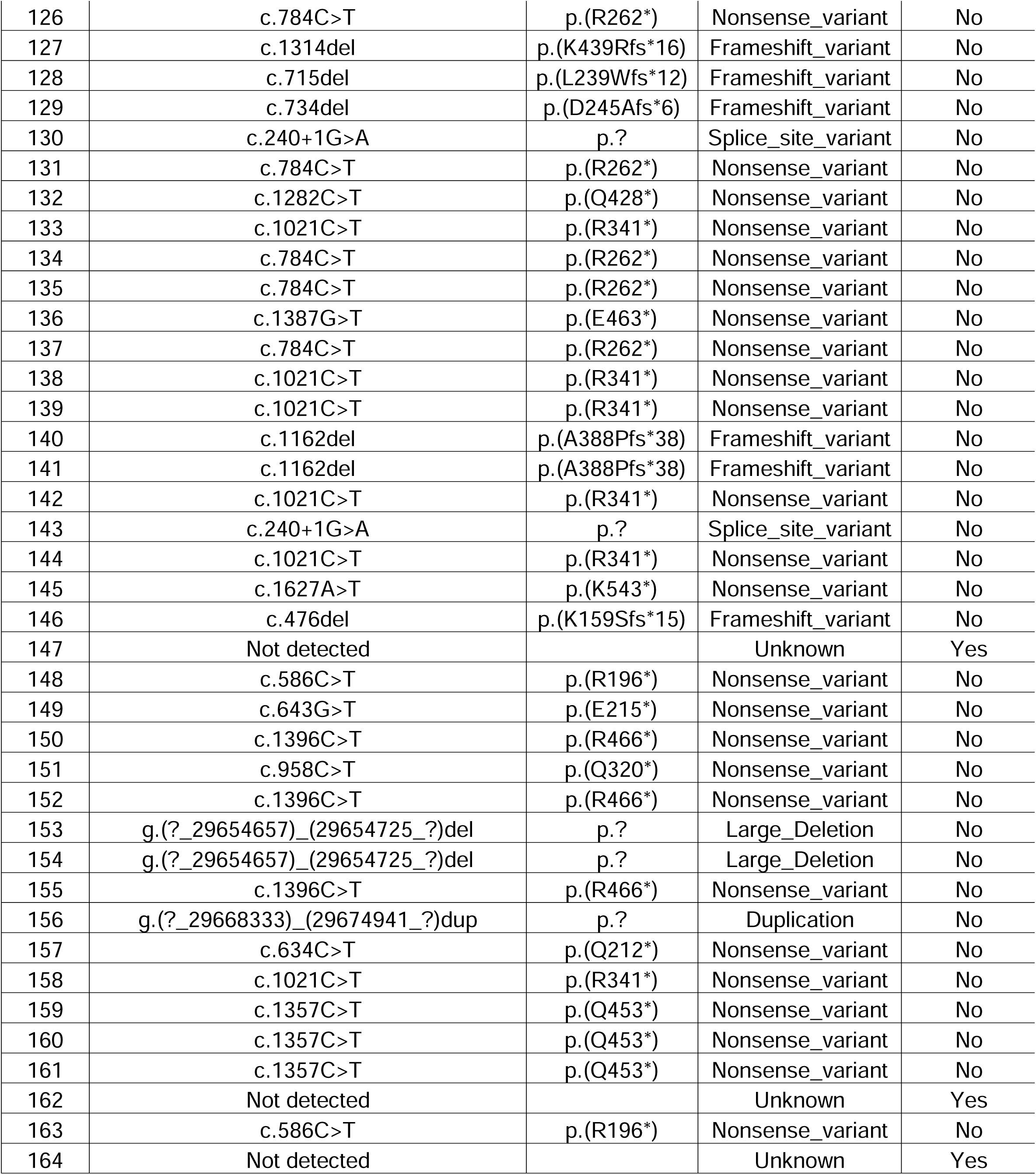
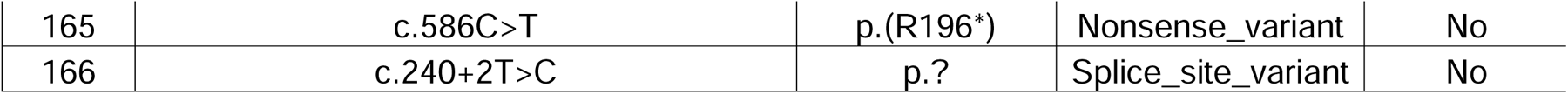
Pathogenic and likely pathogenic *NF2* variants (single nucleotide variants, indels, and copy number variants) identified in the full cohort.

**Sup. Table 2:** Combination of germline and somatic *NF2* variants in our cohort (n=37)

| Patient | Germline *NF2* variant | Somatic *NF2* variant |
| --- | --- | --- |
| 97 | g.(?_29668333)_(29698598_?)del | c.655G>A |
| 119 | c.1030C>T | LOH |
| 119 | c.1030C>T | Not detected |
| 71 | c.193C>T | LOH |
| 3 | Not detected | Not detected |
| 120 | c.1021C>T | LOH |
| 66 | c.1090_1099del | c.105del |
| 72 | c.592C>T | LOH |
| 75 | c.1003G>T | Copy neutral LOH |
| 75 | c.1003G>T | c.361C>T |
| 125 | c.1188dup | LOH |
| 127 | c.1314del | Copy neutral LOH |
| 53 | g.(?_29603633)_(29671948_?)del | Not detected |
| 53 | g.(?_29603633)_(29671948_?)del | c.586C>T |
| 53 | g.(?_29603633)_(29671948_?)del | c.676-1G>T |
| 128 | c.715del | LOH |
| 129 | c.734del | c.592dup |
| 79 | c.1387G>T | LOH |
| 80 | c.1627A>T | LOH |
| 131 | c.784C>T | LOH |
| 18 | c.1030C>T | LOH |
| 18 | c.1030C>T | LOH |
| 82 | c.855del | LOH |
| 135 | c.784C>T | LOH |
| 135 | c.784C>T | Not detected |
| 40 | c.810_810+2dup | LOH |
| 108 | c.447+2T>C | LOH |
| 64 | Not detected | LOH |
| 64 | Not detected | Not detected |
| 87 | c.1341-2A>C | LOH |
| 88 | c.634C>T | LOH |
| 27 | Not detected | LOH |
| 142 | c.1021C>T | LOH |
| 112 | g.(?_29642202)_(29654725_?)del | LOH |
| 30 | Not detected | LOH |
| 30 | Not detected | LOH |
| 116 | c.169C>T | Copy neutral LOH |

## Supplemental figures

**Sup. Fig. 1.**
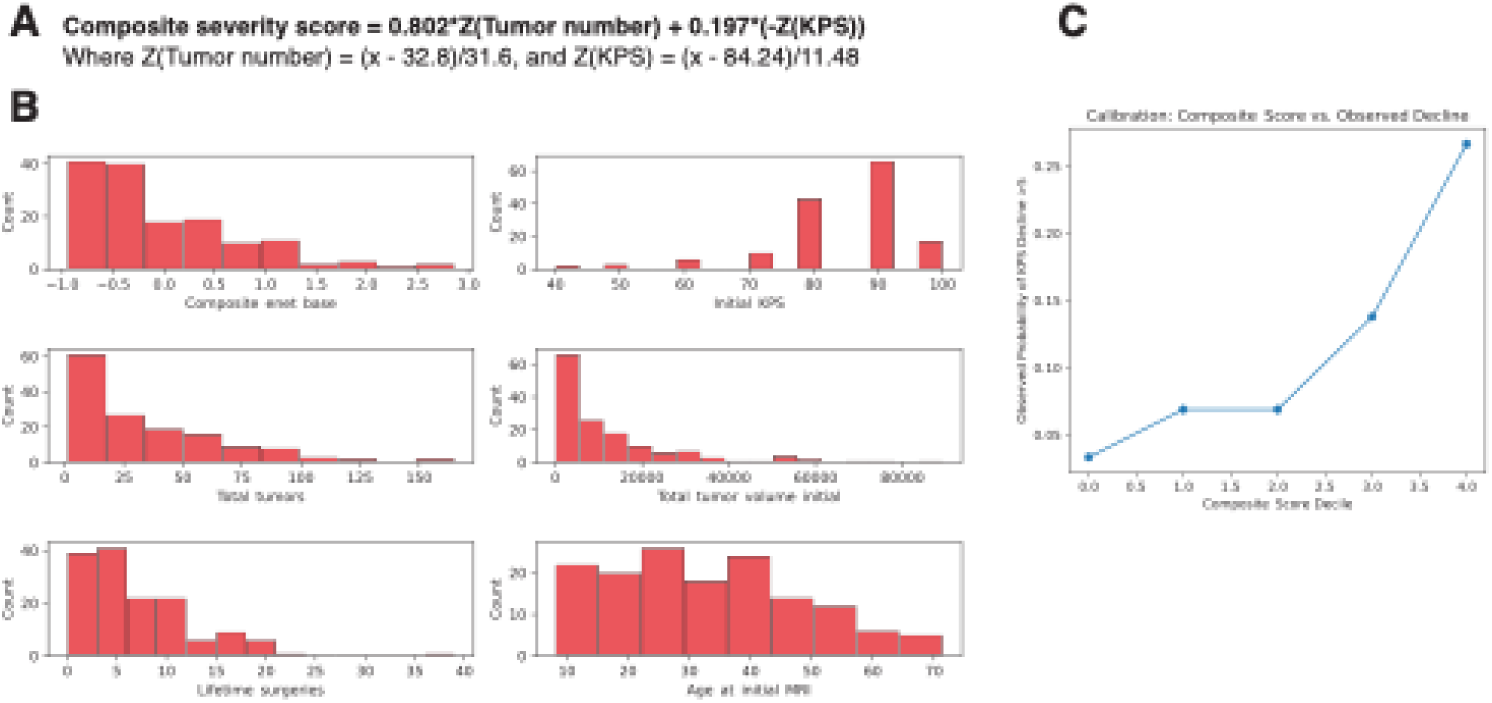
Composite Severity Score (CSS): derivation, distribution, and clinical correlation. **A.** Formula used to calculate the Composite Severity Score (CSS). **B.** Histograms showing the cohort-wide distributions of CSS alongside its five input parameters-initial KPS, total tumor count, total tumor volume, lifetime surgeries, and age at initial MRI-demonstrating that the CSS distribution faithfully reflects the underlying input data. **C.** Monotonic relationship between increasing CSS and stepwise decline in KPS.

**Sup. Fig. 2.**
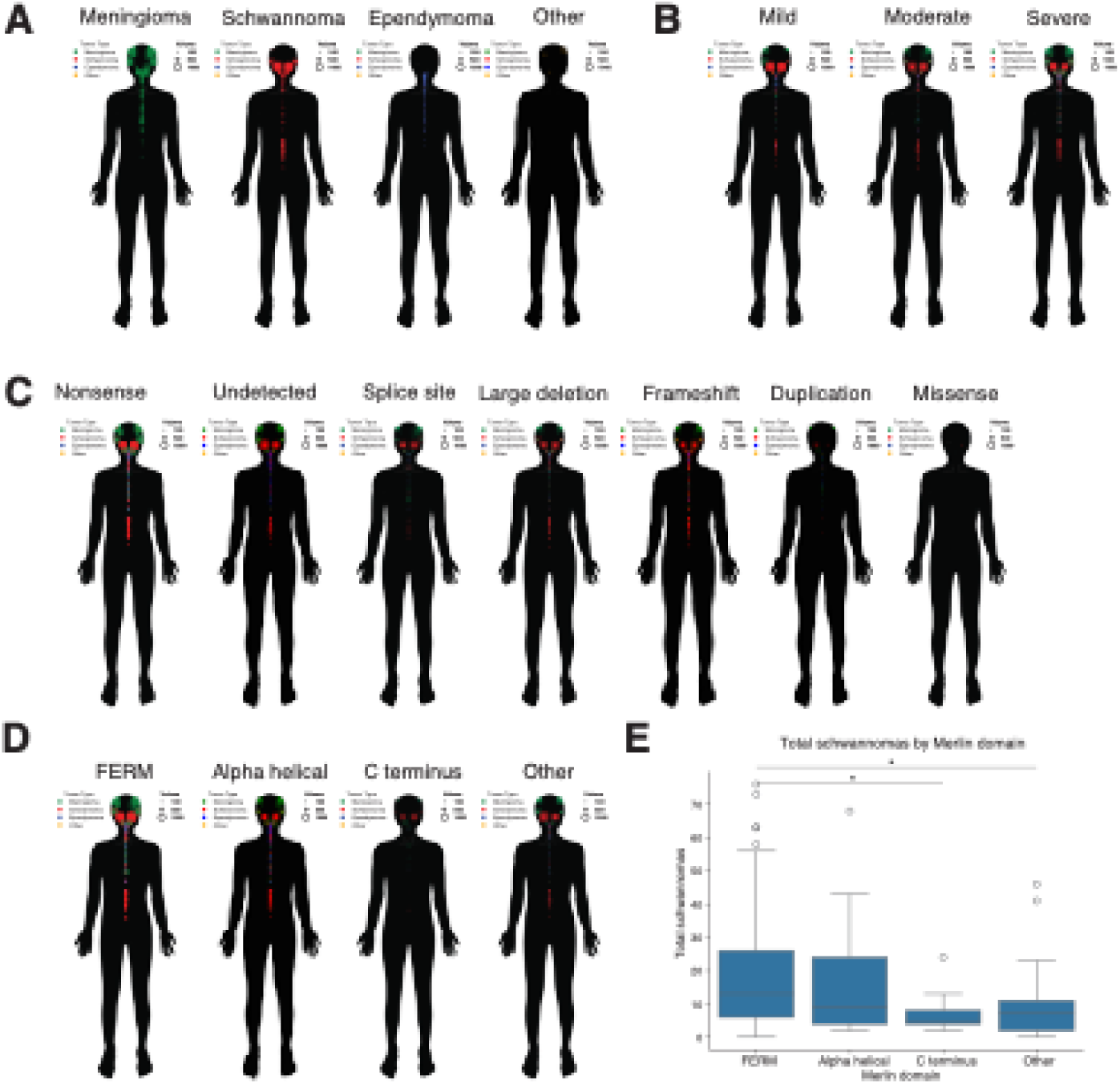
Anatomic distribution of NF2-SWN tumors across severity, variant type, and merlin functional domain. **A.** Anatomic map illustrating the spectrum of tumor types associated with NF2-SWN. **B.** Anatomic map depicting tumor distribution stratified by severity tertile. **C.** Anatomic map depicting tumor distribution stratified by variant type. **D.** Anatomic map depicting tumor distribution stratified by merlin functional domain. **E.** Box-and-whisker plot demonstrating schwannoma incidence by domain, with enrichment observed in the FERM and alpha-helical domains.

**Sup. Fig. 3.**
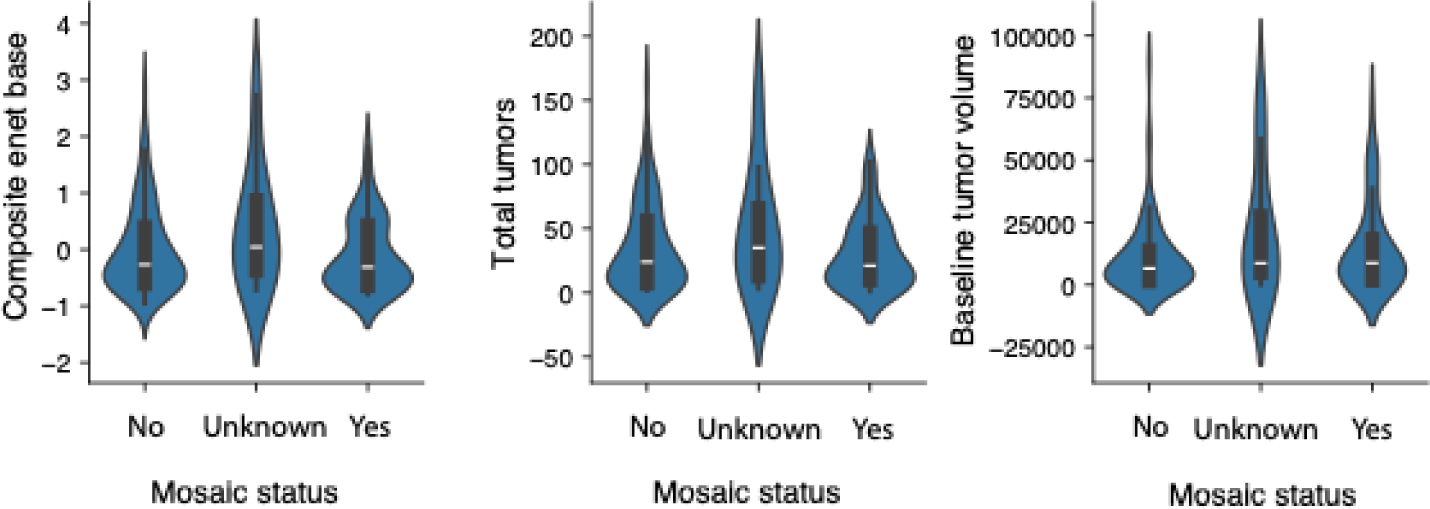
Impact of mosaicism on disease burden in NF2-SWN. Parallel violin plots comparing composite severity score, total tumor count, and total tumor volume between mosaic and non-mosaic NF2-SWN patients.

**Sup. Fig. 4.**
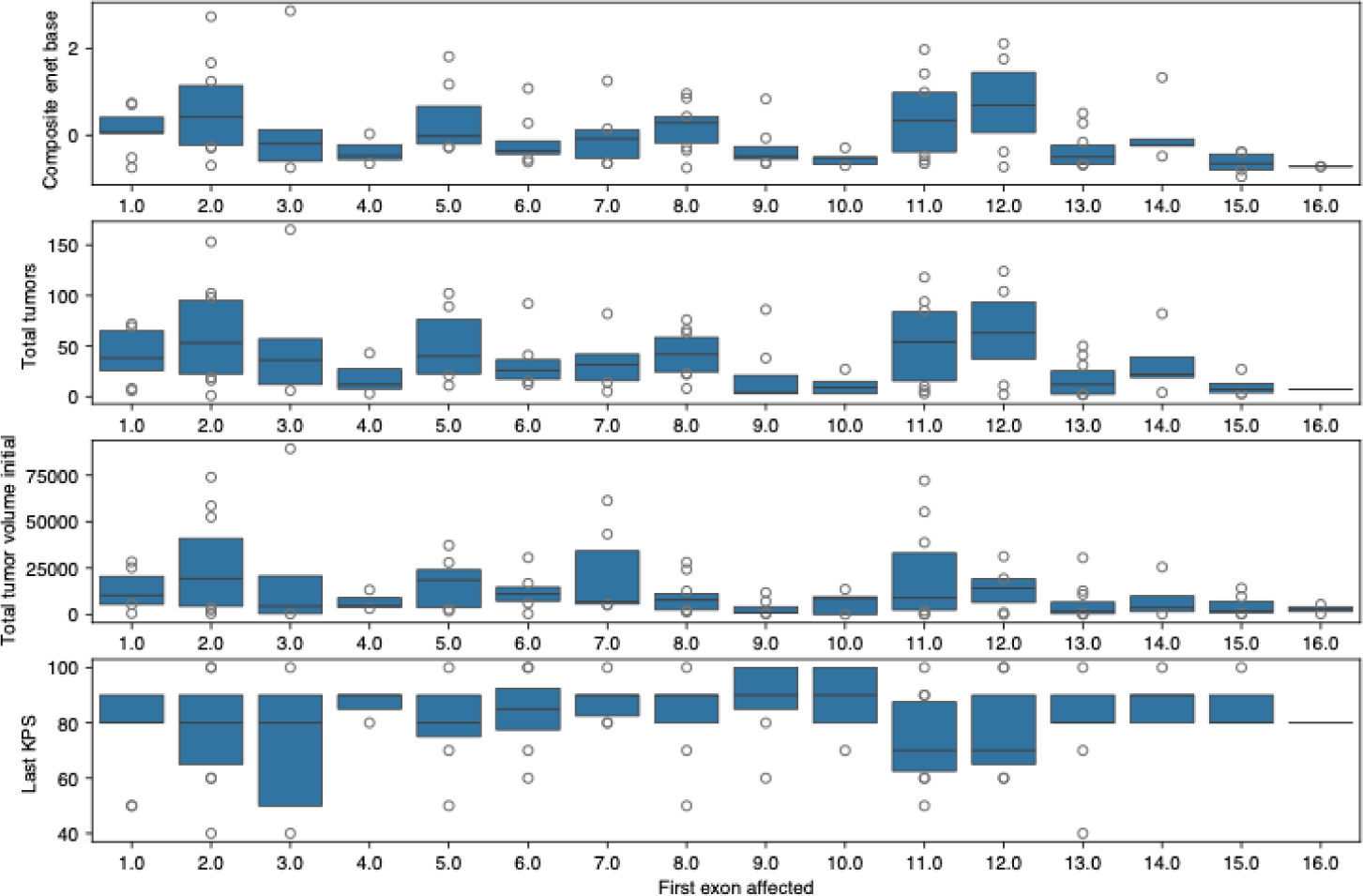
Composite Severity Score captures disease burden more robustly than individual metrics. Stacked bar plots comparing the distribution of NF2-SWN disease severity across CSS, total tumor count, total tumor volume, and KPS, demonstrating the superior consistency of CSS relative to individual measures.

## Notes

### Competing Interest Statement

The authors have declared no competing interest.

### Author Declarations

Combined Neuroscience Institutional Review Board of the National Institutes of Health gave ethical approval of this work

### Summary of Updates

Corrected an error in the author's name.

